# EchoAtlas: A Conversational, Multi-View Vision-Language Foundation Model for Echocardiography Interpretation and Clinical Reasoning

**DOI:** 10.64898/2026.03.14.26348388

**Authors:** Chieh-Ju Chao, Mohammad Asadi, Lavonda Li, Gokul Ramasamy, Nicolo Pecco, Yu-Chiang Wang, Timothy Poterucha, Reza Arsanjani, Garvan Kane, Jae K Oh, Imon Banerjee, Curtis Langlotz, Li Fei-Fei, Ehsan Adeli, Bradley J. Erickson

**Author notes:** **Address of Correspondence:** 200 1st Street SW Rochester, MN 55095, USA Chieh-Ju Chao, MD Assistant Professor of Medicine, Department of Cardiovascular Medicine, Mayo Clinic Twitter: @chiehjuchao1.

## Abstract

Echocardiography is the most widely used cardiac imaging modality, yet artificial intelligence-enabled interpretation remains limited by the inability of existing models to integrate visual assessment, quantitative measurement, and clinical reasoning within a unified framework. Here we present EchoAtlas, the first autoregressive vision-language model developed for echocardiographic interpretation. Trained on over 12.9 million question-answer pairs derived from approximately 2 million echocardiogram videos, EchoAtlas achieves 0.966 accuracy on multiple-choice questions in our internal test set and establishes a new state-of-the-art on the public MIMIC-EchoQA benchmark (0.699 vs. 0.508 previously). EchoAtlas also provides accurate quantitative measurements, segment-level regional wall motion assessment, longitudinal comparison, and diagnostic reasoning across diverse question formats — capabilities not previously demonstrated in this domain. These results highlight the potential of autoregressive vision-language models as a foundation for interactive echocardiographic interpretation, representing an early step toward scalable, auditable artificial intelligence systems in cardiology practice.

## Introduction

Echocardiography (echo) serves as the primary imaging modality in cardiology, utilizing high-temporal-resolution ultrasound video to assess cardiac structure and function^1,2^; however, its interpretation remains labor-intensive and subject to inter-observer variability^3–5^. Over the past decade, advances in artificial intelligence (AI) have demonstrated the potential for automation in echo analysis. While single-task models have achieved promising results in isolated objectives such as ejection fraction estimation and disease classification ^4,6–8^, they exhibit critical limitations: inability to integrate findings across multiple cardiac structures, inflexibility in answering diverse clinical questions beyond pre-defined tasks, and lack of natural language explanations that facilitate clinical understanding and decision-making^9^.

To address these challenges, foundation models with multi-task capabilities are increasingly being explored for echocardiographic analysis^5,10–12^. Recent approaches include models like PanEcho and EchoApex, which focus on direct prediction of multiple specific diseases and quantitative measurements ^11,12^, and vision-language models (VLMs) that integrate visual and textual information for interpretation tasks^5,10^. Among VLMs, Contrastive Language-Image Pretraining (CLIP)-style contrastive models such as EchoCLIP and EchoPrime, trained on more than a million of echocardiogram videos^5,10^, have achieved robust performance on multiple prediction and classification tasks. EchoPrime also demonstrated the potential of interpretation through a retrieval mechanism^5^.

In the broader medical imaging domain, autoregressive VLMs have demonstrated success in general-purpose medical visual question answering (VQA) and report generation^13–16^. These works demonstrate that VQA is an emerging capability for medical VLMs, enabling natural language interaction without constraining clinicians to predefined tasks. Furthermore, autoregressive VLMs provide reasoning capabilities that enhance clinical audit that CLIP-style contrastive models lack^5,10,13^. However, no autoregressive VLM has been developed specifically for echo interpretation. This gap likely reflects the complexity of video-text alignment and the scarcity of publicly available echo datasets suitable for VLM training. The need for precise quantitative measurements in echo has been further considered a potential limiting factor for autoregressive VLMs, which are optimized for language generation rather than structured numerical output. Consequently, progress on echo VQA has remained limited; the current state-of-the-art (SOTA) on the MIMIC-EchoQA benchmark is 50.8% accuracy, achieved by Echo-CoPilot, an agentic system combining EchoPrime and GPT-5.1^17–19^.

In this context, we propose that an autoregressive VLM trained specifically for echo can enable flexible VQA, measurements, longitudinal comparison, and visual-based clinical reasoning in a unified framework. Such a model would represent the first autoregressive VLM designed for echo interpretation, capable of answering diverse clinical questions while providing explicit reasoning that supports transparent and auditable clinical decision-making.

## Results

### Generic Visual Question Answering

We compared the resulting system, EchoAtlas **(Figure 1**), against the base Qwen2.5VL-7B and general medical vision-language models, including OctoMed-7B, MedGemma-4B-IT, on our internal test set (**Table 1**). EchoAtlas variants, EchoAtlas-Temp and EchoAtlas-Joint, developed to assess the feasibility of Template Reporting (see Methods), demonstrated similar performance across these 5 categories. When compared to other models, EchoAtlas demonstrated significantly superior performance across all question formats. For multiple choice questions (MCQ) and closed-ended questions, EchoAtlas achieved 0.966 and 0.943 accuracy, with a significant margin compared to 0.161-0.464 for other models. When stratified by anatomical keywords, EchoAtlas demonstrated robust and balanced performance, achieving MCQ and Close accuracy above 0.9 for nearly all cardiac structures, with a similar significant margin over other comparison models (**Supplementary Figures 1–2**). On Description, Open, Comparison, and Reasoning questions, EchoAtlas also showed marked advantages with RadGraph F1 scores of 0.365–0.785 versus OctoMed’s 0.176-0.212 and MedGemma-4B-IT’s 0.076-0.342 (**Supplementary Figures 3–5**).

**Figure 1.**
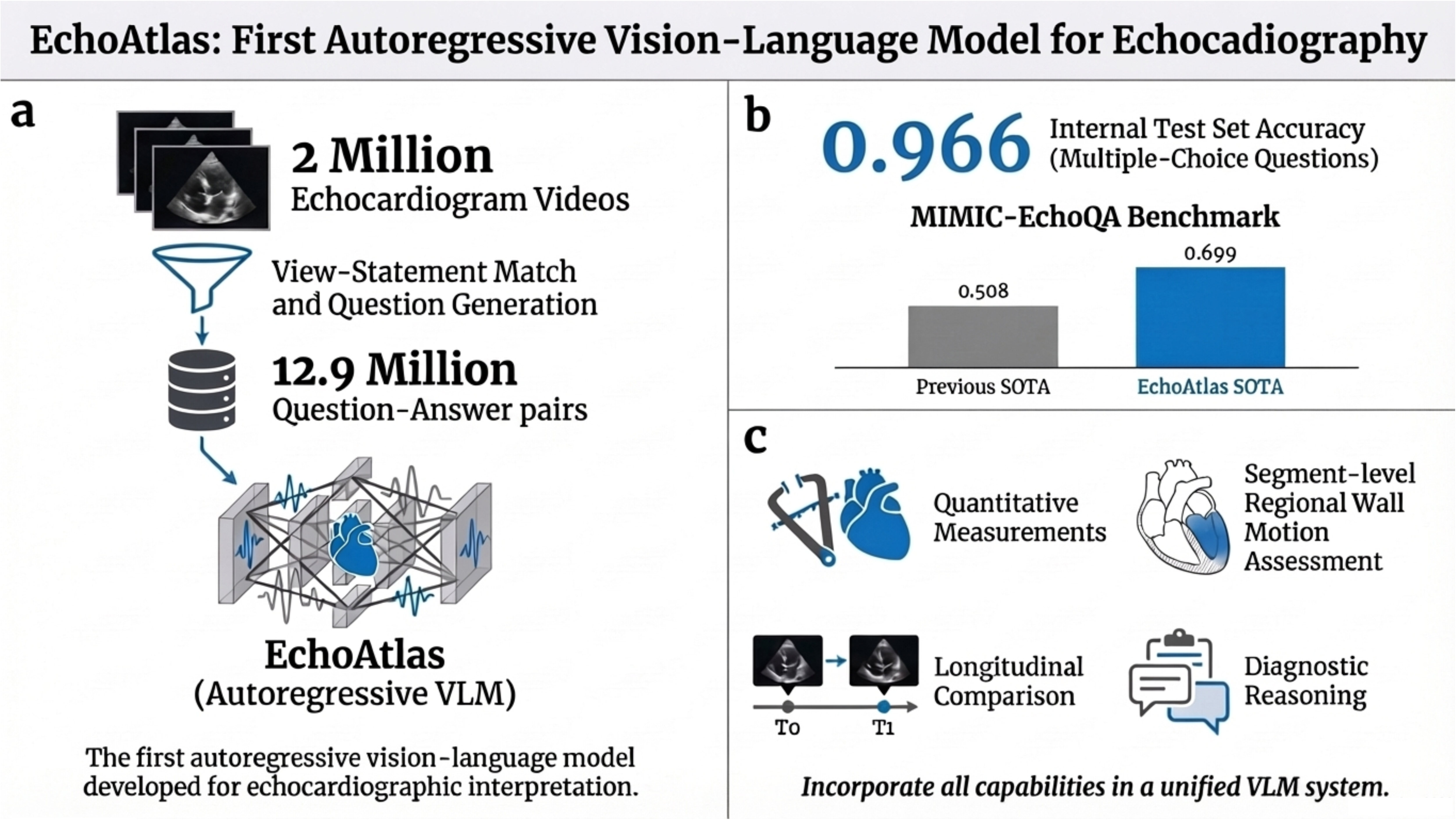
Overview of EchoAtlas Study. Panel a. Development and Performance of EchoAtlas. EchoAtlas is an autoregressive vision-language model (VLM) trained on a large-scale dataset of 12.9 million question-answer pairs corresponding to ∼2 million echocardiographic videos. Panel b. Benchmark performance. The model achieves 0.966 accuracy on internal MCQ test sets and establishes a new state-of-the-art (SOTA) on the public MIMIC-EchoQA benchmark (0.699 accuracy) compared to previous benchmarks (0.508). Panel c. In addition to MCQ, EchoAtlas also demonstrates robust performance across clinical tasks, including quantitative measurements, segment-level regional wall motion assessment, longitudinal study comparison, and complex diagnostic reasoning.

**Table 1.** Model’s Performance on Internal Test Set.

| Question Format | Metric | EchoAtlas | EchoAtlas-Temp | EchoAtlas-Joint | OctoMed-7B | MedGemma-4 B-IT | Qwen2.5VL-7B | N. of examples | N. of tokens |
| --- | --- | --- | --- | --- | --- | --- | --- | --- | --- |
| MCQ | Accuracy | 0.966 (0.964 - 0.967)*†‡ | 0.966 (0.965 - 0.967)*†‡ | 0.967 (0.965 - 0.968)*†‡ | 0.161 (0.158 - 0.164) | 0.422 (0.418 - 0.426) | 0.363 (0.359 - 0.367) | 63752 | 5.4 ± 5.7 |
| Close | Accuracy | 0.943 (0.940 - 0.945)*†‡ | 0.943 (0.940 - 0.945)*†‡ | 0.943 (0.941 - 0.946)*†‡ | 0.409 (0.403 - 0.414) | 0.464 (0.459 - 0.470) | 0.375 (0.370 - 0.380) | 34146 | 3.4 ± 6.1 |
| Description | RadGraph F1 | 0.785 (0.782 - 0.787)*†‡ | 0.786 (0.783 - 0.788)*†‡ | 0.784 (0.781 - 0.786)*†‡ | 0.212 (0.210 - 0.215) | 0.342 (0.339 - 0.345) | 0.332 (0.330 - 0.335) | 32452 | 18.4 ± 6.7 |
| Open | RadGraph F1 | 0.703 (0.701 - 0.705)*†‡ | 0.703 (0.701 - 0.705)*†‡ | 0.702 (0.700 - 0.704)*†‡ | 0.228 (0.226 - 0.229) | 0.293 (0.291 - 0.294) | 0.290 (0.289 - 0.291) | 39639 | 38.2 ± 10.4 |
| Comparison | RadGraph F1 | 0.581 (0.551 - 0.612)*†‡ | 0.572 (0.542 - 0.604)*†‡ | 0.584 (0.557 - 0.617)*†‡ | 0.070 (0.067 - 0.074) | 0.282 (0.261 - 0.305) | 0.151 (0.141 - 0.160) | 518 | 14.0 ± 5.4 |
| Reasoning | RadGraph F1 | 0.365 (0.352 - 0.379)*†‡ | 0.367 (0.354 - 0.380)*†‡ | 0.372 (0.359 - 0.385)*†‡ | 0.176 (0.168 - 0.183) | 0.076 (0.072 - 0.079) | 0.115 (0.110 - 0.120) | 558 | 428.3 ± 51.4 |
| Template Report | Accuracy | N/A | 0.783 (0.763 - 0.803)*†‡ | 0.787 (0.767 - 0.807)*†‡ | 0.016 (0.011 - 0.023) | 0.478 (0.453 - 0.502) | 0.467 (0.442 - 0.492) | 1801 | 2.5 ± 1.1 |

Performance was expressed as mean (95%CI). \*p value < 0.05 when compared to OctoMed-7B, †p value < 0.05 when compared to MedGemma, ‡p value < 0.05 when compared to Qwen2.5VL-7B-Instruct. Ground truth correction: An initial mapping error affected 22 (4.2%) temporal comparisons in the Comparison Category. All reported metrics reflect corrected ground truth.

OctoMed’s frequent refusal to answer when uncertain likely stems from its instruction-tuning, which prioritizes clinical safety over response rate. The performance gap was consistent across all question formats.

### Measurements

The model demonstrated strong overall performance on measurement prediction tasks across 27,391 numerical (R²=0.897, MAE=6.73) and 3,478 wall motion categorical measurements (3-class accuracy 0.745). For numerical predictions, the model achieved notably high R² values for several key measurements, including LV cardiac index (R² = 0.999, MAE = 0.02), right ventricular systolic pressure (R² = 0.845, MAE = 2.485), and Left Atrial Maximum Volume by both the two-chamber (R² = 0.768, MAE = 8.00) and four-chamber (R² = 0.913, MAE = 5.80) methods, and Left Ventricular End Diastolic Volume (R² = 0.886, MAE = 10.94) and end systolic volume (R² = 0.829, MAE = 9.07), indicating reliable quantitative estimation of these cardiac parameters. Ejection fraction predictions by both two-chamber and four-chamber views achieved R² values of 0.77 and 0.46, respectively, with MAEs of 2.54% and 4.68% (**Figure 2a and 2b**). For 3-class (normal, hypokinesis, akinesis) categorical wall motion scoring across 16 segments, the model achieved accuracies between 0.586 and 0.892. Basal antero-lateral and mid anterior wall segments showed the highest performance (0.883 and 0.892, respectively), while inferior and infero-lateral segments were comparatively more challenging (0.586 –0.704). In binary classification (normal vs. abnormal), our model also achieved robust performance with a positive predictive value of 77.1% and a negative predictive value of 85.3%, with details summarized in **Figure 2c**. Compared to other models, EchoAtlas-Temp achieved comparable performance, while EchoAtlas-Joint exhibited worse performance despite being trained on fewer measurement tasks (**Figure 2a and 2b**). MedGemma failed to produce meaningful predictions for most numerical measurements, except for 2-chamber left atrial volume (R² = 0.798, MAE = 8.194); all remaining measurements yielded low or negative R² values. Its wall motion score accuracy was 0.542. OctoMed-7B also failed across nearly all measurement tasks (**Figure 2**).

**Figure 2a and 2b.**
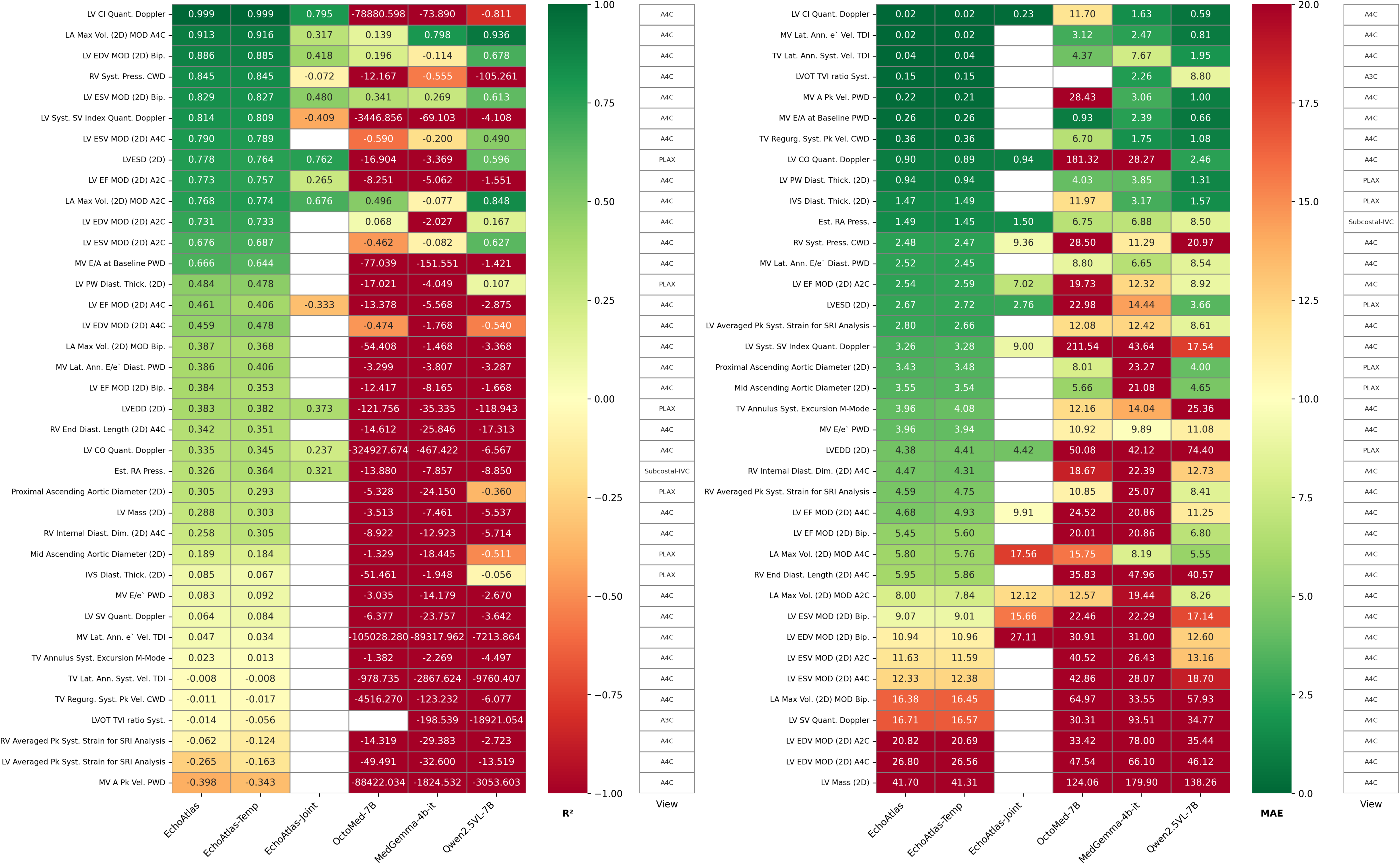
. Model performance for echocardiographic measurements assessed by R² and MAE. Heatmaps showing R² values (a) and MAE values (b) across multiple cardiac measurements predicted by different models. Higher R² values and lower MAE values (green) indicate better prediction accuracy. The model achieved notably high R² values for LV cardiac index (0.999), right ventricular systolic pressure (0.845), left atrial maximum volume by four-chamber view (0.913), and left ventricular end diastolic volume (0.886), with corresponding low MAEs of 0.02, 2.485, 5.80, and 10.94, respectively, demonstrating reliable quantitative estimation of key cardiac parameters. Ejection fraction predictions showed R² values of 0.77 (two-chamber) and 0.46 (four-chamber) with MAEs of 2.54% and 4.68%, respectively.

**Figure 2c.**
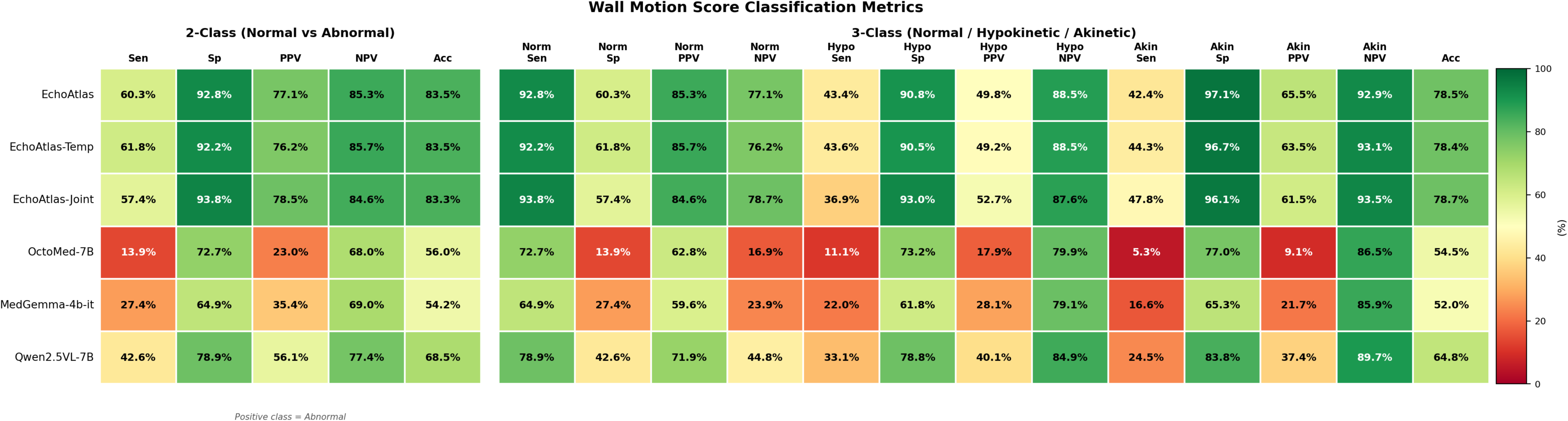
Wall motion score classification performance across models. Heatmap comparing six vision-language models on 3,478 echocardiographic segments. Left panel: binary (normal vs. abnormal); right panel: 3-class (normal/hypokinetic/akinetic). Unparseable predictions were counted as failures across all metrics. EchoAtlas variants achieved binary accuracy of 0.833–0.835, PPV of 0.771–0.786, and NPV of 0.845–0.853 for abnormality detection, representing 15–20 percentage point improvements over the base Qwen2.5VL-7B model. OctoMed-7B and MedGemma-4B-IT showed substantially lower performance (binary accuracy 0.560 and 0.542; PPV 0.230 and 0.354; NPV 0.680 and 0.690). Sen, sensitivity; Sp, specificity; PPV, positive predictive value; NPV, negative predictive value; Acc, overall accuracy.

### Diagnostic Reasoning and Subsection Analysis

EchoAtlas and variants demonstrated the best performance compared to other models in clinical reasoning tasks (**Table 1**). Note that inter-model comparisons used overall reasoning content, as other models lack structured subsection generation. EchoAtlas underwent additional subheading-level analysis for granular assessment. Disease-specific performance varied both in overall scores and across reasoning subsections. The top-performing diseases were aortic stenosis (range: 0.267–0.435 across subsections, n=140), mitral stenosis (range: 0.218–0.391, n=46), and mitral regurgitation (range: 0.201–0.331, n=86). The poorest performance was observed in aortic dissection (range: 0.060–0.275, n=4) and cardiac tamponade (range: 0.135–0.242, n=8). By reasoning subsections, visual observations from echocardiogram videos achieved the highest average RadGraph F1 score (mean: 0.306, range: 0.231–0.397 across diseases), followed by application of diagnostic criteria (mean: 0.288, range: 0.208–0.435) and integration (mean: 0.248, range: 0.159–0.343). Hemodynamic interpretation demonstrated the poorest performance (mean: 0.192, range: 0.066–0.280), along with motion pattern analysis (mean: 0.194, range: 0.060–0.267) (**Figure 3a**).

In the 50 reasoning cases for experts’ review, reasoning exhibited near-perfect relevance (5.00±0.00) and coherence (5.00±0.00), with strong completeness (4.96 ± 0.14), while correctness showed moderate variability (4.00 ± 0.81). The structured six-subsection format ensured comprehensive coverage from visual observations through clinical integration.

Performance varied by disease category (**Figure 3b**), with mitral regurgitation achieving the highest correctness (4.33) and cardiac tamponade the lowest (3.00). The model’s reasoning quality primarily depended on initial diagnostic accuracy, with correct diagnoses leading to appropriate downstream reasoning. There are 8% of cases had insufficient image quality (judged by either expert). The agreement between the two annotators were moderate (agreement rate 86.4%, Cohen’s kappa= 0.553).

### Template Reporting

Across the Template Reporting multiple-choice questions, the two EchoAtlas variants (EchoAtlas-Temp and EchoAtlas-Joint) substantially outperformed all baselines (**Table 1**). EchoAtlas-Joint achieved the highest overall accuracy (0.787, 95% CI: 0.767–0.807), followed closely by EchoAtlas-Temp (0.783, 95% CI: 0.763–0.803), with particular strength on detection tasks such as intracardiac mass/thrombus (1.000), TV prosthesis (0.983), and pericardial effusion (0.949), while performance was more modest on complex tasks such as mitral leaflet morphology (0.427) and 4-grade MR/TR severity (∼0.55). MedGemma-4B-IT and Qwen2.5VL-7B showed intermediate accuracy (0.478, 95% CI: 0.453–0.502, and 0.467, 95% CI: 0.442–0.492, respectively). OctoMed-7B was essentially non-functional on this task, achieving near-zero accuracy across virtually all 26 templates (overall accuracy 0.016, 95% CI: 0.011–0.023) with non-parseable responses. For the two continuous measurement questions, EchoAtlas-Joint showed modest but positive R² for LVEF (0.294) and RVSP (0.131), while EchoAtlas-Temp achieved lower values (R²=0.137 and 0.036, respectively); the baseline models either lacked sufficient valid numeric responses or produced negative R² values, indicating no meaningful predictive ability for quantitative measurements (**Supplementary Figure 6**).

### External Validation on MIMIC-EchoQA

We evaluated EchoAtlas’ external performance on the public MIMIC-EchoQA benchmark (n=622 questions) and compared it against MedGemma-4B-IT, OctoMed-7B, QwenVL2.5, and adapted EchoPrime, with Echo-CoPilot representing the current SOTA. EchoAtlas achieved a 0.699 accuracy (95% CI: 0.662-0.734), significantly outperforming the prior SOTA, Echo-CoPilot (0.508), adapted EchoPrime (0.481; 95% CI: 0.442-0.520), and other models (all p< 0.001) (**Figure 4a**). Notably, Echo-CoPilot is an agentic framework leveraging EchoPrime and GPT-5.1 as its core components^18^; our adapted EchoPrime implementation achieved comparable performance to the original system. Both EchoPrime and Echo-CoPilot performed similarly to other generic medical VLMs like OctoMed-7B and MedGemma-4B-IT (**Figure 4a**). The performance gap of EchoAtlas is consistent when stratified by structures and views, leading in almost all categories (**Figures 4b and 4c**). While not shown in the figure, EchoAtlas-Temp and EchoAtlas-Joint had worse accuracies, 0.665 (95% CI: 0.628-0.701) and 0.333 (95% CI: 0.314-0.365), respectively. The performance of other baseline models on this benchmark, including Qwen2.5-VL-7B, Qwen2.5-VL-72B, GPT-4o, Qwen2-VL-2B-biomed, and Qwen2-VL-7B-biomed, ranged from 35%-45%^17,18^. Minor performance variations were observed when re-evaluating some of these models, likely attributable to differences in prompt design.

**Figure 3a.**
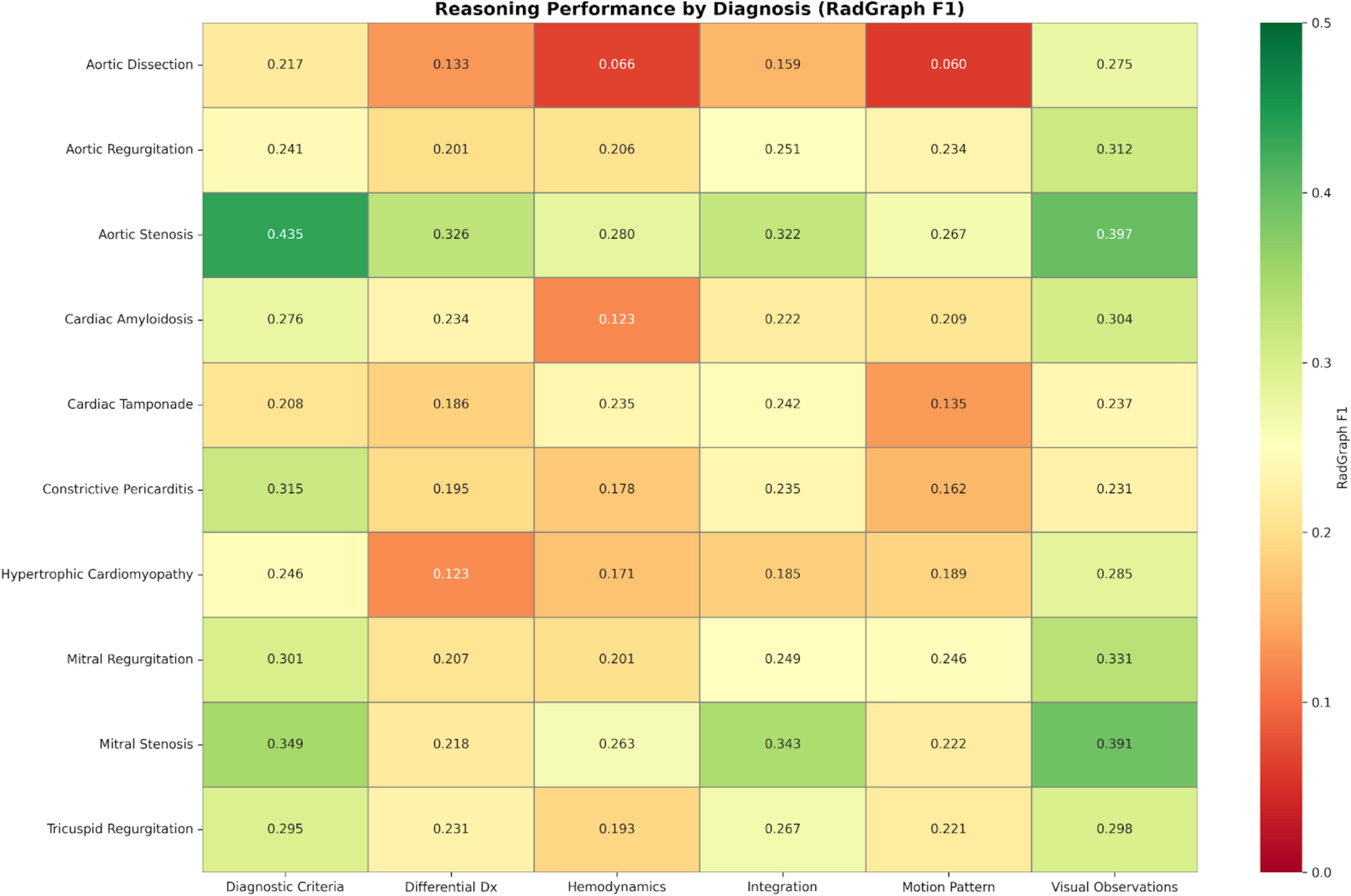
Long-form reasoning performance across diagnostic subsections assessed by RadGraph F1 score. Heatmap showing model performance on different reasoning subsections (Diagnostic Criteria, Differential Diagnosis, Hemodynamics, Integration, Motion Pattern, and Visual Observations) across ten cardiac diagnoses. Higher RadGraph F1 scores (green, scale 0-0.5 for better visualization) indicate better alignment between model-generated and reference reports. EchoAtlas demonstrated robust long-form reasoning capabilities, with Visual Observations achieving the highest scores across all diagnoses (mean F1: 0.275-0.397), followed by Diagnostic Criteria (0.208-0.435) and Differential Diagnosis (0.123-0.326). Aortic Stenosis showed the strongest overall performance across subsections, while more complex diagnoses like hypertrophic cardiomyopathy and cardiac tamponade presented greater reasoning challenges.

**Figure 3b.**
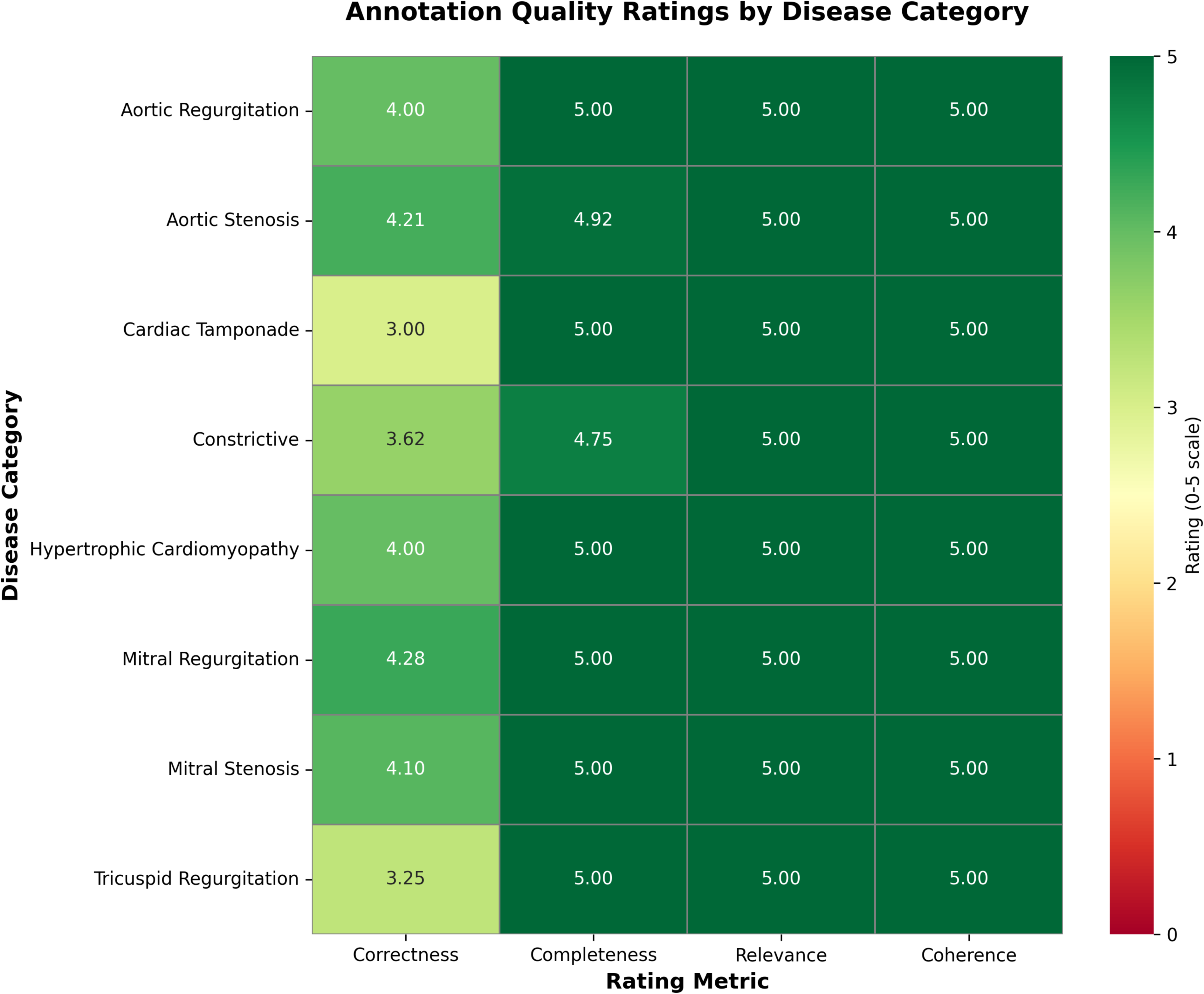
Expert evaluation of reasoning quality across diagnostic categories. Heatmap showing model performance on different reasoning quality metrics (correctness, completeness, relevance, and coherence) across 8 diseases in a subset of randomly selected cases. Reasoning demonstrated near-perfect relevance (5.00±0.00) and coherence (5.00±0.00), strong completeness (4.94±0.25), and moderate correctness (3.87±0.95). Performance varied by disease category, with mitral regurgitation achieving the highest correctness (4.28) and cardiac tamponade the lowest (3.00).

**Figure 4.**
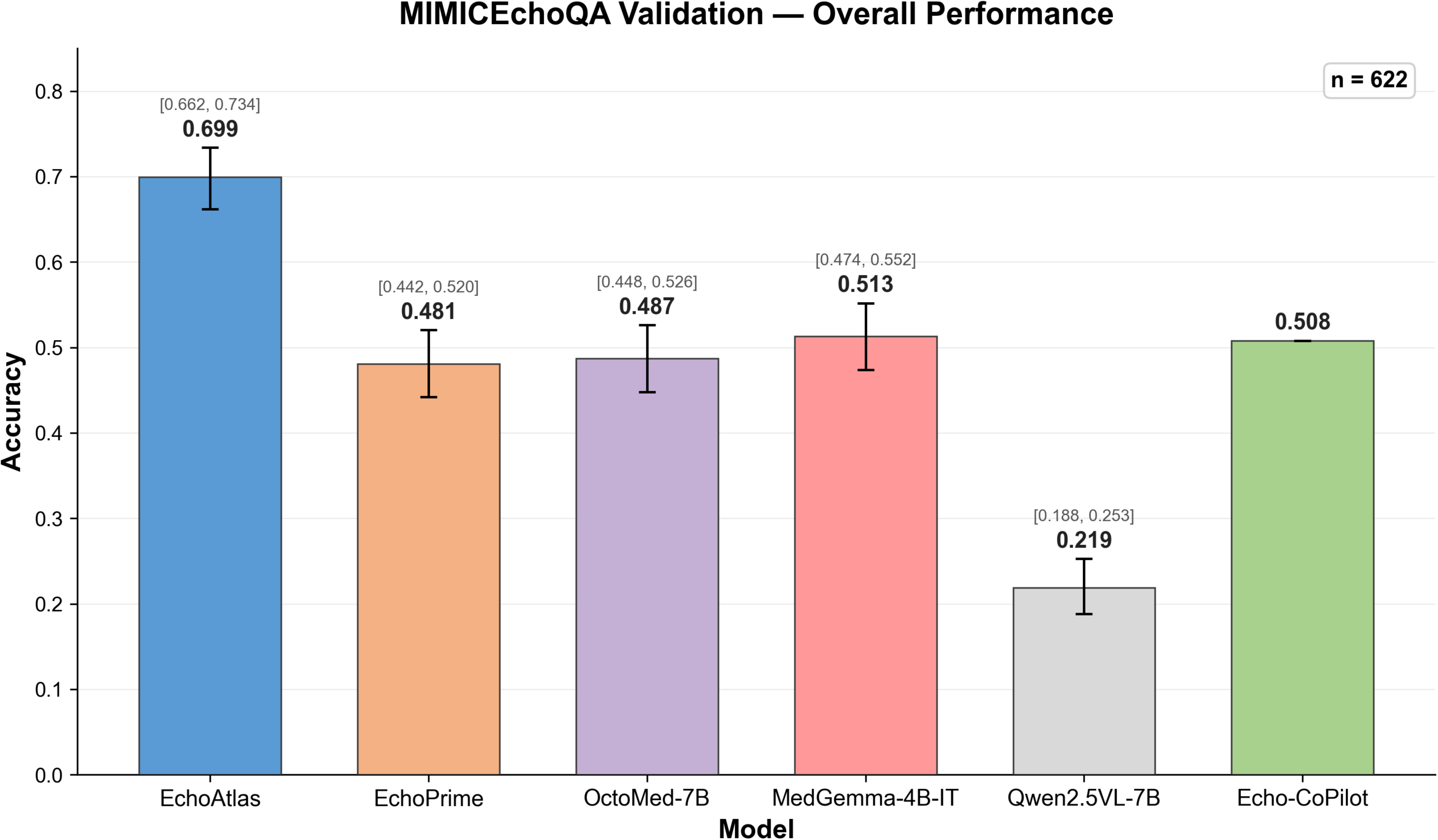
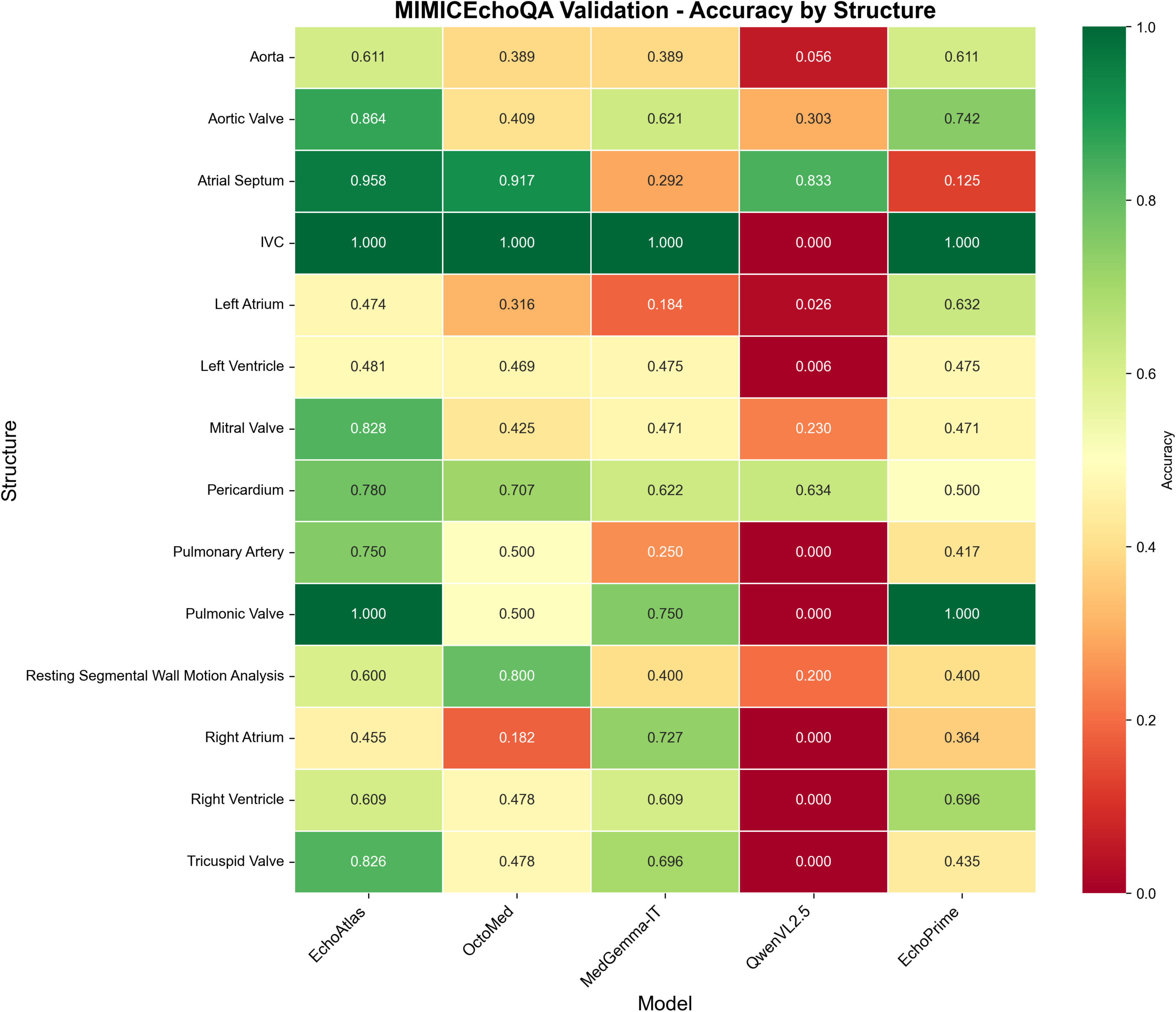
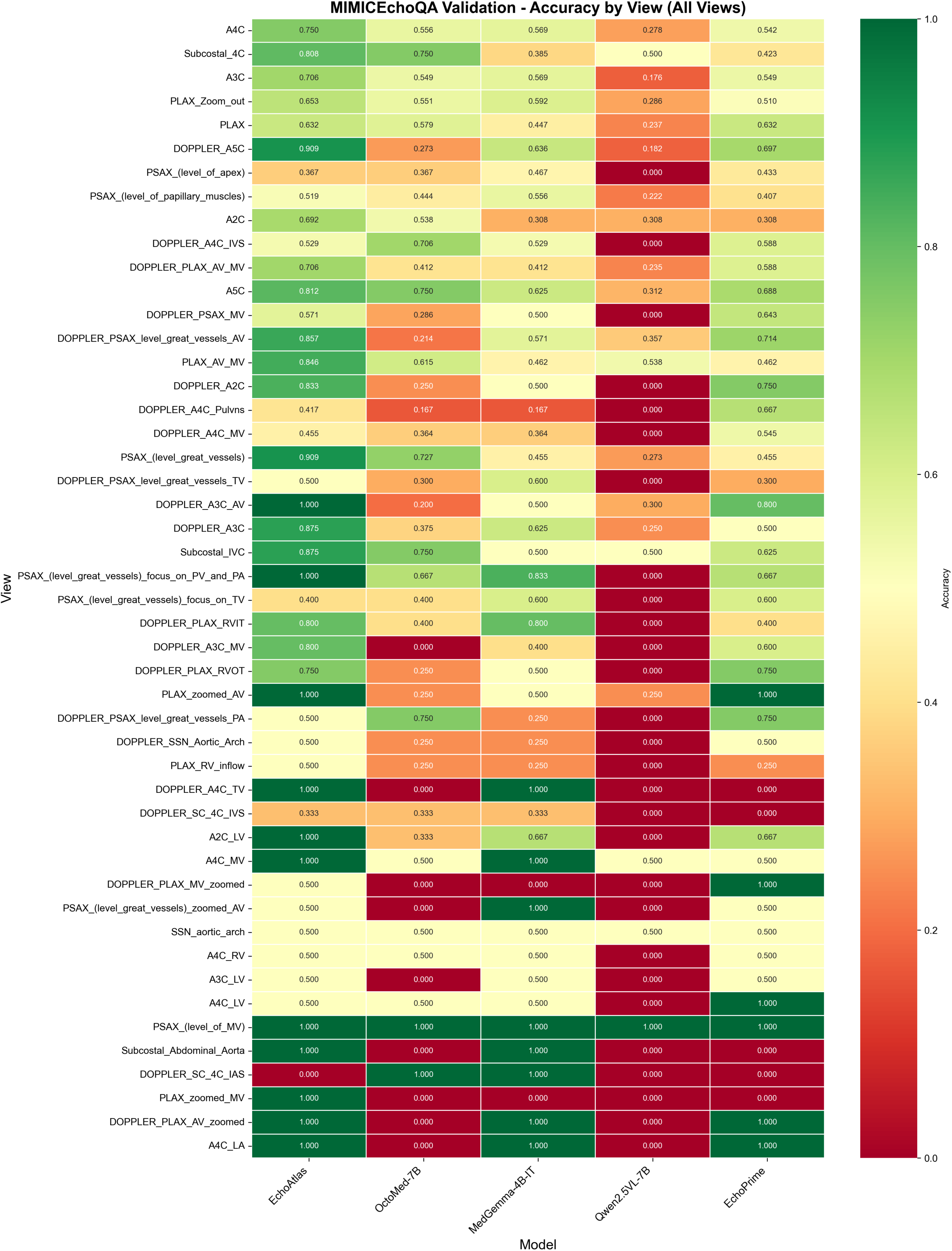
**EchoAtlas vs other models on MIMIC-EchoQA Benchmark**. **Panel a.** Comparison of MCQ accuracy between EchoAtlas and other models. Error bars and bracketed values indicate the 95% CI for models. Echo-CoPilot (SOTA) is shown to represent the published accuracy from literature without CIs. All models were evaluated on the same test set (n=622 questions). EchoAtlas achieves 0.699 accuracy [95% CI: 0.662-0.734], significantly outperforming the current SOTA performance of Echo-CoPilot (50.8%) and other models. EchoPrime and Echo-CoPilot have similar performance to other general-purpose medical VLMs. **Panels b and c.** Model performance stratified by structure and view, respectively. EchoAtlas showed superior performance in nearly all categories except for the left atrium and right ventricle.

After expert review of all 187 errors of EchoAtlas by two independent raters, an average of 39.6% represented true model failures. The majority of errors were attributable to ambiguous ground truth at subjective severity boundaries (39.3%), label noise in the benchmark (10.2%), view-question mismatches (5.6%), or poor image quality (5.3%).

Both raters noted that the lack of multi-view data was the primary cause of the high ratio of ambiguous ratings. Inter-rater agreement was fair (agreement rate 47.6%, Cohen’s κ = 0.25).

## Discussion

In this work, we present EchoAtlas, the first conversational VLM for echocardiographic interpretation. Trained on over 12.6 million properly designed visual questions spanning approximately 2 million echocardiogram videos, we developed a unified echo VQA interpretation system capable of performing visual assessments, quantitative measurements, longitudinal comparison, and clinical reasoning within a conversational framework. Unlike prior approaches that either focus solely on predictions or combine predictions with textual report retrieval^5,11,12^, EchoAtlas integrates all interpretation capabilities into a single autoregressive model, with all tasks formatted as VQA. This unified framework enables seamless expansion of clinical capabilities through dataset curation alone, allowing rapid adaptation to evolving reporting standards without architectural modification. Critically, prior works rely on study-level aggregation that obscures view-specific findings^5,11^, whereas our view-level approach enables direct interpretation and audit of individual videos. EchoAtlas demonstrated robust performance, achieving an overall 0.966 MCQ accuracy, and establishing new SOTA performance on the external MIMIC-EchoQA benchmark (accuracy 0.699)—an absolute 19.1-point improvement over prior SOTA^18^.

EchoAtlas demonstrated robust performance on the internal test set for echocardiographic VQA tasks, outperforming generic medical VLMs by over 40% (**Table 1**), underscoring the value of domain-specific training at scale. Its strong accuracy on MCQs, including disease severity grading, suggests potential for integration into echo interpretation workflows with minimal hallucination risk under structured MCQ constraints. Beyond MCQs, EchoAtlas is the first model in the field to demonstrate general conversational VQA capability spanning diverse question formats, including measurements, longitudinal comparison, and diagnostic reasoning, marking a significant step toward applying autoregressive VLMs in this field.

Among quantitative parameters, EchoAtlas achieved R² ≥ 0.7 for 29% and R² ≥ 0.3 for 63% of all 39 measurements (**Figure 2a**). Of the remaining 14 parameters with R² < 0.3, more than half were Doppler-based measurements evaluated on their most relevant views (A4C or PLAX). This likely reflects the absence of Doppler spectral images in our training data. In regional wall motion abnormality detection, a task with known inter-observer variability, EchoAtlas demonstrated strong performance at the segment level, providing finer spatial resolution than the wall/territory or global level assessments in prior works^5,11,20–22^, while achieving comparable or superior diagnostic performance (**Figure 2c**). This approach demonstrates that VLMs can perform accurate measurement prediction by treating numerical values and units as natural language tokens, enabling seamless integration of quantitative and qualitative assessments within a unified model. Unlike traditional multi-task architectures requiring task-specific prediction heads, this formulation allows flexible expansion to new measurements through dataset design alone, without architectural modifications, representing a key advantage for adapting to evolving clinical reporting standards.

For free-text questions, we observed anticipated superior performance in the Open and Description categories compared to the Comparison and Reasoning categories, reflecting the latter’s increased complexity and response length requirements (**Table 1**). EchoAtlas demonstrated robust long-form reasoning capabilities, with visual observation achieving the highest scores across all subsections, followed by differential diagnosis and diagnostic criteria, which indicates solid visual understanding and relevant echocardiographic knowledge (**Figure 3**). The relatively weaker performance on hemodynamic analysis was expected, given its complexity and the absence of Doppler spectrum images or measurements (e.g., velocity, gradient) typically required for accurate hemodynamic interpretation. Notably, natural language processing metrics penalized valid paraphrasing, though RadGraph F1 > 0.3 typically reflects clinically satisfactory content in prior studies^14,23^. Performance on reasoning tasks may be further improved by increasing the proportion of reasoning training examples, which currently represent 0.2% of the dataset due to the requirement for specific cases. Nevertheless, EchoAtlas demonstrated superior reasoning performance compared to general-purpose medical VLMs (**Table 1**).

In terms of generalization, EchoAtlas established a new SOTA performance, with 69.9% accuracy on MIMIC-EchoQA^19^ (19.1-point improvement over prior SOTA, Echo-Copilot’s 50.8%)^18^, demonstrating robust generalization across institutions and temporal robustness to data. Notably, the MIMIC-EchoQA benchmark^19^ uses a 1-question-1-video setting that does not fully reflect clinical practice, which often requires synthesizing information across multiple views and measurements. This explains the high rate (39.6%) of ambiguous ground truth in expert error analysis, as cardiology experts often need additional information to make judgments. Future echo VQA benchmarks should consider incorporating multi-view, multi-measurement frameworks.

Our evaluation revealed a trade-off between template reporting and conversational VQA. While joint training from initialization (EchoAtlas-Joint) marginally improved template accuracy (0.787 vs. 0.783 for EchoAtlas-Temp’s staged training), it triggered catastrophic degradation in internal measurement and external VQA performance, dropping from 0.699 to 0.333 in accuracy. These failures may stem from multiple factors. First, to avoid out-of-memory error, our template data organization (1-2 videos per required view) differs from other VQA tasks (3-5 videos per required view), potentially limiting the depth of visual information available to the model.

Second, enforcing a uniform template structure across all studies may introduce artificial constraints that conflict with the adaptive QA capability required for diverse question types. Whether these challenges reflect dataset design limitations, template format constraints, or genuine architectural incompatibility remains an open question requiring further investigation.

Our comparison between EchoPrime^5^ and EchoAtlas on the MIMIC-EchoQA benchmark^17,19^ illustrates a fundamental trade-off in echo vision-language model design. CLIP-style retrieval-augmented approaches like EchoPrime excel at comprehensive interpretation with complete studies but can suffer performance degradation with partial inputs (**Figure 4**)^5^. Also, combining predictions with retrieval-based reporting introduces inconsistencies when predicted findings contradict retrieved template text (**Supplementary Figure 7**)^5^. Furthermore, while EchoPrime’s prediction components generalized well^5^, retrieval-based reporting inherently limits cross-institutional generalizability by assuming training corpus templates match deployment scenarios. Critically, this architecture precludes conversational capability and restricts interpretability to study-level attention information^5^, limiting clinical audit capabilities.

Unlike CLIP-style or multi-task predictive models^5,11,12^, EchoAtlas offers greater flexibility by interpreting relevant partial inputs independently while collectively covering the full study, maintaining data fidelity. Its view-level design enables auditable, computationally efficient interpretations on demand, and directly addresses the multi-video-to-one-report alignment challenge inherent in study-level aggregation approaches. As a trade-off, view-selection sensitivity was observed in measurement performance, suggesting view-task alignment as a promising direction for future improvement. Autoregressive architectures also enable further enhancement through reinforcement learning for clinical reasoning^14,24,25^ and potentially numerical precision, with hallucination risk mitigatable through structured query formats. We envision EchoAtlas as an interactive co-pilot for echo interpretation, with carefully designed prospective human-AI collaborative protocols essential to realizing its full clinical potential.

This study has several limitations. The model was trained on single-institution data from Mayo Clinic and validated on MIMIC-EchoQA from Beth Israel Deaconess Medical Center^19^; however, both are academic medical centers, and generalizability to community hospitals with different patient populations and practice patterns remains uncertain. The retrospective design requires prospective validation to assess real-world clinical utility. Similar to other prior works, the scope was limited to 2D grayscale and color Doppler videos of echo studies, while other excluded acquisitions could potentially improve the model performance^5,11^. Our measurement capabilities cover key clinical parameters, but not all possible echocardiographic quantifications. Future iterations incorporating multiple templates tailored to specific clinical contexts could further enhance reporting accuracy and clinical applicability. Finally, although larger VLM architectures are available, our model’s strong performance (>90% accuracy on MCQ and other tasks) suggests diminishing returns from scaling, particularly given the computational efficiency tradeoffs for clinical deployment.

## Methods

### Datasets

Mayo Clinic Echo Data: For adult patients with a research consent, we identified all adult transthoracic echo studies from 1/1/2023-12/31/2023 with matched study and report from Mayo Clinic. All images were retrieved in DICOM format.

**Echocardiogram Preprocessing and View Classification.** DICOM files were categorized based on frame count and color Doppler presence. We specifically identified the following four categories: multi-frame with/without color Doppler, and single-frame with/without color Doppler. Multi-frame videos underwent preprocessing adapted from existing protocols to isolate the ultrasound sector and remove ECG tracings via morphological masking and convex hull operations. Frames were cropped (10% edge trimming), resized to 224×224 pixels, and exported as MJPEG AVI files at 30 fps^5^. For view classification, we employed a DINOv2 vision transformer (ViT-B/14 with register tokens) pre-trained via self-supervised learning^26^, fine-tuned with a custom head (768→512→16 dimensions with dropout layers) to predict 16 standard echocardiographic views (A2C, A3C, A4C, PLAX, PSAX variants, subcostal views, and color Doppler views) with a 99% AUC. During inference, 11 frames were uniformly sampled from each video using linear interpolation. Per-frame predictions were aggregated via majority voting among predictions with confidence ≥0.7, returning the consensus view or null if no confident predictions emerged. Quality control filtering subsequently excluded videos with classification confidence <0.7 (no view assigned) or mismatches between predicted Doppler status and DICOM metadata, removing 19.1% of the dataset (388,640 videos).

**Visual Question Generation.** Following the above process, each of the 16 views was matched to the corresponding statements in the structured report as designated by an expert cardiologist. This view-statement mapping ensured that questions were answerable based on the visible image content. For example, statements regarding the aortic valve were excluded from apical four-chamber views. Conditions potentially visualizable across multiple views, such as hypertrophic cardiomyopathy and cardiac amyloidosis, were assigned to all relevant views. Subsequently, we employed a HIPAA-compliant gpt-4o endpoint licensed to Mayo Clinic (OpenAI, Inc., API version: 2024-10-21) to generate visual questions with paired answers based on the structured report statements. The model was prompted to generate questions under the premise that it could observe the echocardiogram images, though it only had access to the textual reports. The generation parameters were set to max_tokens=6000, temperature=0.1, and top_p=1.0 to ensure deterministic and concise outputs. To capture the diversity of clinical queries, eight formats of questions were generated in two stages. Initially, questions were created across four basic formats (multiple-choice, closed-ended, description, and open-ended) with random view-level assignment. Importantly, MCQ questions were designed to evaluate multi-class severity grading (e.g., differentiating mild, moderate, and severe mitral regurgitation or left ventricular enlargement) rather than simpler binary classification tasks (e.g., presence vs. absence of severe disease). Subsequently, additional task-specific categories were introduced: quantitative measurements, longitudinal comparisons, clinical reasoning, and template reporting. The MCQ prompt template is in the Supplemental file as an example (**Supplementary Figure 9**).

For MCQ, close, open, and description questions, questions were also organized into three hierarchical tiers based on complexity: single-view structural assessments (tier 1, average 2.3 videos per entry), multi-view color Doppler evaluations (tier 2, average 2.7 videos per entry), and multi-view structural assessments with diagnostic integration (tier 3, average 5.7 videos per entry). This tiered approach enables evaluation of both isolated single-view interpretation capabilities and the model’s ability to synthesize information across multiple views, reflecting the clinical workflow where cardiologists integrate findings from complementary echocardiographic perspectives to formulate comprehensive diagnoses.

All splits were performed at the patient level to prevent data leakage. After quality control filtering, the dataset of 62,726 patients was divided into training (61,260 patients, 97.7%), validation (493 patients, 0.8%), and test (973 patients, 1.6%) sets (**Table 2**). This allocation prioritized computational efficiency while ensuring robust evaluation, with the test set alone comprising over 200,000 questions across all task categories (**Supplementary Table 1**). Each patient contains an average of 31.0 videos, reflecting the multi-view nature of clinical echocardiographic interpretation.

**Table 2.**
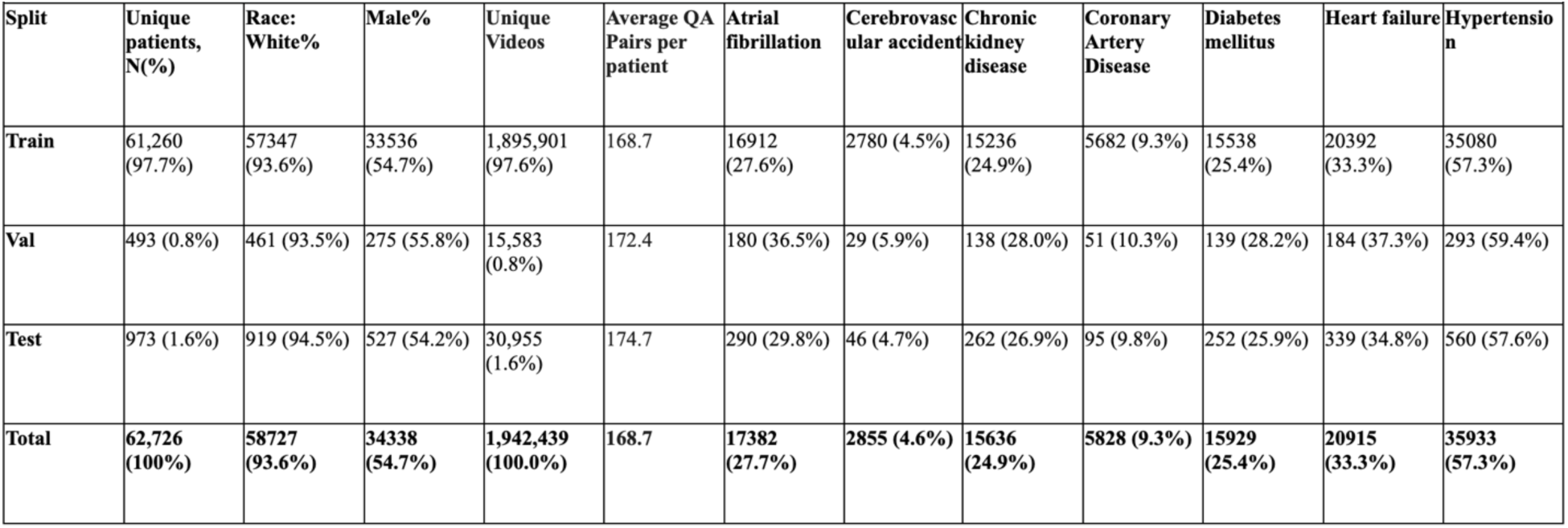
Dataset characteristics.

For Measurement, Comparison, Template Report, and Reasoning questions, multiple videos were included to enable synthetic interpretation across views. **Measurement questions** encompassed parameters across two categories: (1) 38 numerical measurements, including dimensional, volumetric, and Doppler-derived values, with Doppler measurements assigned to their most relevant views; and (2) 16 categorical assessments of regional wall motion (normal/hypokinesis/akinesis). **Comparison questions** were generated for patients with at least two echo studies in the study year, comparing identical findings (e.g., left ventricular ejection fraction) between studies. **Reasoning questions** required a Chain-of-Thought style, long-form responses structured into six clinical reasoning subsections to prevent metric inflation from template repetition while enabling fine-grained analysis: (1) Visual observations from echocardiogram videos, (2) Motion pattern analysis, (3) Hemodynamic interpretation, (4) Differential considerations, (5) Integration and application of diagnostic criteria, and (6) Diagnostic summary. **Template Report questions** employed a generic, standardized 29-question template (**Supplementary Table 2**) organized into sections corresponding to the most relevant views. The same gpt-4o endpoint was used to generate answers most consistent with the original report text for each section. Through this process, we curated a large-scale echo VQA dataset comprising 12.9 million QA pairs, and about 2 million unique echocardiogram videos across 62,726 unique patients (**Table 1**).

**Model selection**. Among open-source autoregressive VLMs, we evaluated candidate architectures based on their performance on established medical image understanding and video reasoning benchmarks. Qwen2.5VL-7B demonstrated superior capabilities in temporally structured visual inputs and medical domain tasks compared to Qwen2VL-7B and InternVL 3.5^27–29^, and was selected as the backbone for EchoAtlas. We adopted the Low-Rank Adaptation (LoRA)^30^ approach to mitigate catastrophic forgetting of the base model’s general vision-language capabilities while accommodating computational resource constraints. Hyperparameter optimization was conducted using a small training subset to identify the optimal configuration. Supervised fine-tuning was implemented using the LLaMA-Factory framework (https://github.com/hiyouga/LLaMA-Factory).

**Staged Training Strategy for Multi-Task Evaluation.** To rigorously assess whether clinical template reporting can be integrated with visual question-answering without performance degradation, we employed a controlled, staged training approach. We first trained EchoAtlas exclusively on VQA tasks (all task types excluding Template Report) to establish baseline performance on conversational interpretation. Subsequently, we continued training this checkpoint for one additional epoch on the Template Report subset to create EchoAtlas-Temp, isolating template generation capabilities through targeted fine-tuning. Finally, we trained EchoAtlas-Joint from initialization with both VQA and Template Report data combined throughout training, incorporating the 13 best-performing numerical measurement tasks identified from EchoAtlas to optimize multi-task learning efficiency. This experimental design enables direct attribution of performance changes to multi-task integration by comparing: (1) baseline VQA-only performance, (2) template-only specialization, and (3) joint multi-task capabilities, thereby isolating the effects of task integration from other confounding factors.

EchoAtlas and variants were trained with the following hyperparameters: LoRA rank of 16, per-device training batch size of 2, gradient accumulation steps of 8, learning rate of 1.0×10⁻⁴, 1 training epoch (2 epochs for EchoAtlas-Temp), cosine learning rate scheduler, and warmup ratio of 0.1. Training was performed on four 80GB NVIDIA A100 GPUs over approximately 190 hours (760 GPU-hours).

**Model Evaluation.** We report the performance on the Mayo internal hold-out test set. As no other published autoregressive medical VLMs capable of interpreting echocardiographic studies currently exist, we compared the model with the base Qwen2.5VL-7B base model, OctoMed-7B, and MedGemma-IT-4B^13,14,29^ as the representations of general-purpose medical VLMs, to assess the value of domain-specific training over general medical AI capabilities across different architectures. Importantly, MedGemma only supports static images rather than videos; therefore, only a single frame from each echocardiogram video was extracted for evaluation with these models.

**Reasoning Quality Analysis**. To assess EchoAtlas generated reasoning, two independent cardiology experts evaluated 50 cases, stratified by disease and randomly selected, across four dimensions established in prior studies: correctness, completeness, relevance, and coherence^23,31,32^. Each dimension was scored on a 5-point Likert scale (1=poor, 5=excellent). These metrics were specifically operationalized to identify common AI failure modes, including hallucinations (false positives) via the correctness metric and missingness (false negatives) via completeness (**Supplementary Table 3**).

**External Validation on MIMIC-EchoQA Benchmark.** MIMIC-EchoQA is a benchmark dataset for echocardiographic VQA derived from the publicly available MIMIC-IV-ECHO database, comprising studies collected at Beth Israel Deaconess Medical Center between 2012–2019^19^. The dataset pairs echocardiogram videos with question-answer pairs generated from cardiologist reports, filtered for view-appropriate content, and validated by board-certified cardiologists. The final dataset contains 622 four-option multiple-choice questions, each aligned with a single echocardiogram video in a 1:1 ratio^17,19^.

On the MIMIC-EchoQA dataset, we benchmarked EchoAtlas against the base Qwen2.5VL-7B model, OctoMed-7B, and MedGemma-IT 4B^13,14,29^. We also explored the potential of EchoPrime^5^ for this VQA format by adapting its architecture to allow single-frame inference, providing a comparison between autoregressive and retrieval-based approaches on this unseen public benchmark.

**EchoPrime Adaptation.** EchoPrime was originally designed for multi-video echocardiogram interpretation, where it synthesizes information across all views in a comprehensive TTE study to generate structured reports via retrieval-augmented generation. To enable comparison on MIMIC-EchoQA, which pairs single videos with targeted questions, we modified the published version of the EchoPrime script to accept single-video input for metrics prediction and retrieval reporting. Then, similar to Echo-Copilot’s approach^18^, the output of EchoPrime is fed into GPT-5 (2024-12-01-preview), with instructions to extract the answer to the sample’s multiple-choice question from the predicted values and the report, as detailed in **Supplementary Figure 9**. We acknowledge this represents an out-of-distribution use case for EchoPrime; however, this comparison reflects a clinically relevant scenario where targeted questions must be answered from individual video clips, a common occurrence in point-of-care settings, focused re-reviews, or when complete studies are unavailable.

**Error analysis.** To distinguish genuine model failures from dataset quality issues, we conducted systematic error analysis on all incorrect EchoAtlas predictions. Two board-certified cardiologists reviewed each error case, examining the input videos, model predictions, and ground truth labels. Errors were categorized into five classes: true model errors, ambiguous ground truth, label noise, view mismatches, and poor image quality (**Supplementary Table 4**).

### Statistical analysis

Following prior VQA works^33^, for multiple-choice questions (MCQs and template reporting) and closed-ended questions, model performance was assessed by accuracy. For Open-ended, Descriptive, Comparison, and Reasoning questions, model performance was assessed using RadGraph F1 to evaluate the correctness of anatomical and pathological entities and their relationships^34^. Numerical measurements were evaluated using R-squared and mean absolute error (MAE). Categorical measurements (16-segment wall motion scores) were assessed by sensitivity (recall), specificity, positive predictive value (precision), negative predictive value, and accuracy in 3-class (normal, hypokinesis, akinesis) and 2-class (normal versus abnormal) settings. For question types beyond Reasoning, Template Reporting, and Measurements, which utilize fixed-format templates, we stratified responses by clinical keywords (e.g., left ventricle, aortic valve stenosis) extracted from the questions to enable more fine-grained performance analysis. For expert error analysis, inter-rater agreement was assessed using agreement rate and Cohen’s kappa, with final scores averaged across the two raters.

For all metrics, we used nonparametric bootstrapping (n = 1,000 replicates) to estimate 95% confidence intervals when appropriate. Observed differences in performance between pairs of models were tested for statistical significance using a two-sided paired permutation test (n = 1,000 permutations), with the null hypothesis being that there is no difference in the performance of the two models. In each permutation, paired prediction outcomes for the two models were randomly swapped to obtain a new difference in model performance. The p-value corresponds to the proportion of permuted differences with an absolute value greater than or equal to the observed difference. Statistical significance was defined as p < 0.05 for all comparisons.

### Ethical review and approval

All the studies have been performed in accordance with the Declaration of Helsinki. The use of the Mayo Clinic dataset was approved by the institutional review board (protocol#22-010944); only patients providing informed consent for minimal-risk retrospective studies were included, thus the requirement for additional informed consent was waived.

## Data Availability

The Mayo Clinic data set is not publicly available due to patient privacy considerations; however, it can be obtained from the corresponding author upon reasonable request and IRB approval. MIMIC-EchoQA benchmark is a public dataset available at PhysioNet https://doi.org/10.13026/rndk-4s36.

## Data Availability

https://doi.org/10.13026/rndk-4s36

## Acknowledgment

CJC is supported by NIH/NHLBI K08 (HL175205-01A1), and grants from the Mayo Clinic, including the MC-AIR Seedling Award, the CV Prospective Award from the Department of Cardiovascular Medicine, and the AI/ML Enablement Award from the Center for Digital Health. MA is supported by the Amazon AI PhD Fellowship and the Stanford HAI Graduate Fellowship. The other authors declare no relevant funding for this work.

## Author Contributions

C.J.C.: Conceptualization, methodology, software, validation, data curation, formal analysis, writing – original draft, visualization.

M.A.: Methodology, software., writing – original draft.

L.L.: Validation, data curation, writing – original draft, visualization. G.M.: Data curation, software.

N.P.: Data curation, software. Y.C.W.: Data curation.

T.P.: Data curation, writing – review and editing. R.A.: Resources, writing – review and editing.

G.C.K.: Resources, writing – review and editing. J.K.O.: Resources, writing – review and editing.

I.B.: Methodology, resources, writing – review, and editing.

F.F.L.: Methodology, resources, writing – review and editing, supervision. C.P.L.: Resources, writing – review and editing, supervision.

E.A.: Conceptualization, methodology, resources, writing – review, and editing. B.J.E.: Conceptualization, Resources, writing – review and editing, supervision. All authors have read and approved the manuscript.

## Abbreviations

Echo: Echocardiography
LLM: Large Language Model
LV: Left Ventricle
MAE: Mean Absolute Error
MCQ: Multiple-Choice Question
NLP: Natural Language Processing
PPV: Positive Predictive Value
NPV: Negative Predictive Value
SOTA: State-Of-The-Art
VLM: Vision-Language Model

---

**Supplementary Figure 1.**
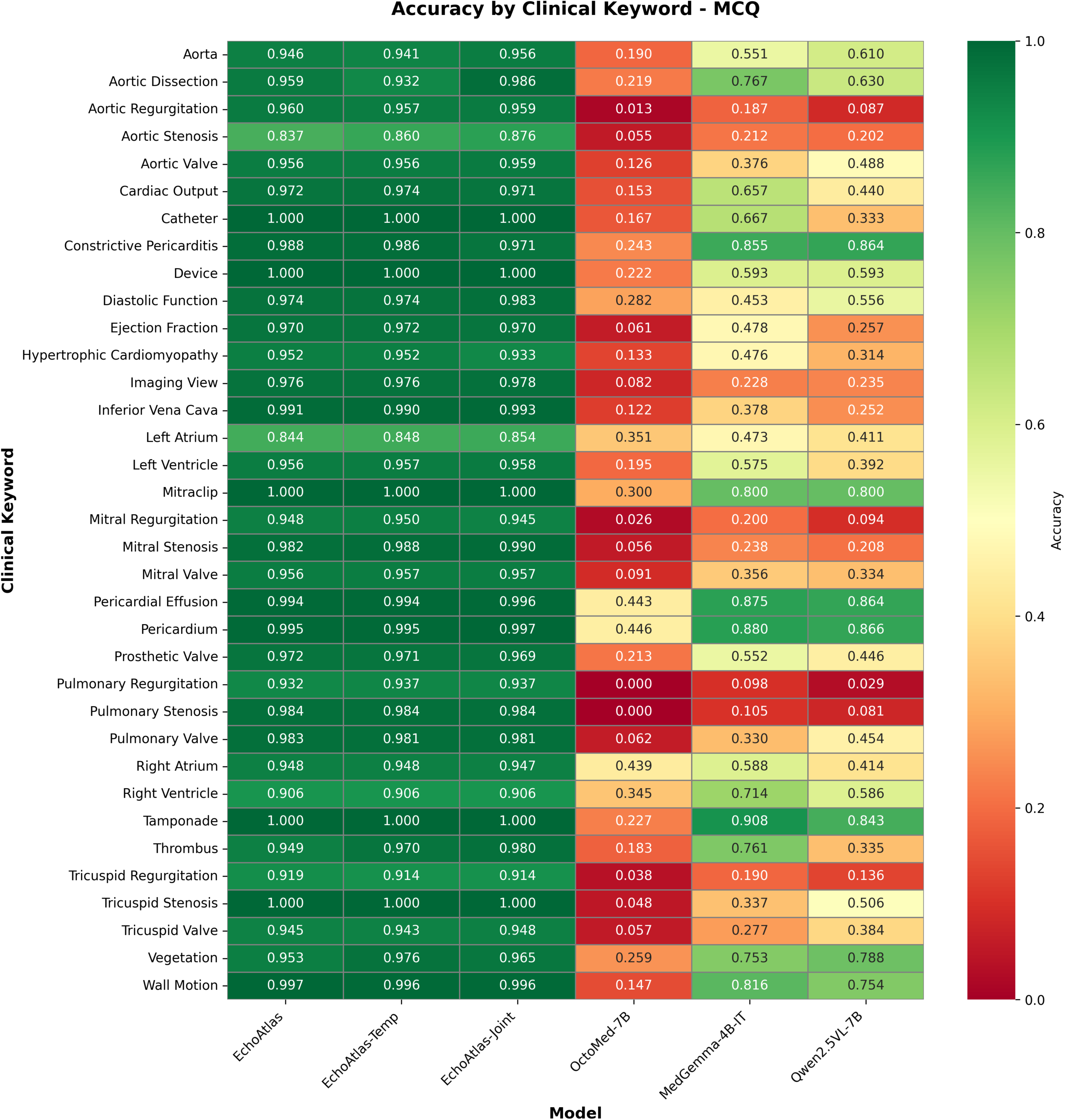
Model performance on multiple-choice questions stratified by clinical keyword. Heatmap displaying accuracy values (color scale 0–1; green indicates higher performance). EchoAtlas achieved above 0.9 accuracy across most categories, except for aortic stenosis (0.837) and left atrium (0.844), and outperformed all comparison models (OctoMed-7B, MedGemma-4B-IT, and Qwen2.5VL-7B) in every category.

**Supplementary Figure 2.**
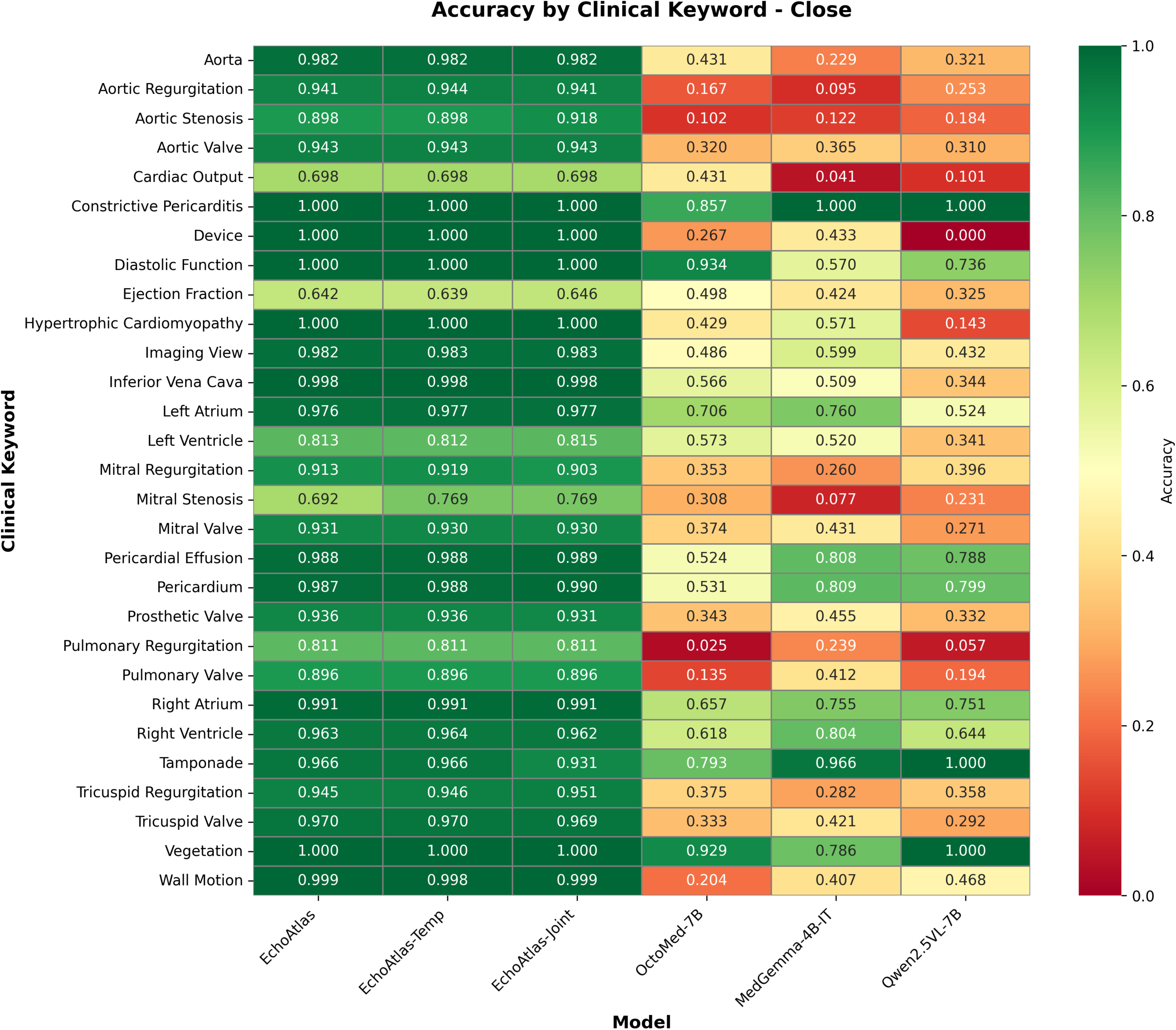
Model performance on closed-ended questions stratified by clinical keyword. Heatmap displaying accuracy values (color scale 0–1; green indicates higher performance). EchoAtlas achieved above 0.9 accuracy across a majority of categories and outperformed all comparison models (OctoMed-7B, MedGemma-4B-IT, and Qwen2.5VL-7B) in every category, except for a tie in constrictive pericarditis.

**Supplementary Figure 3.**
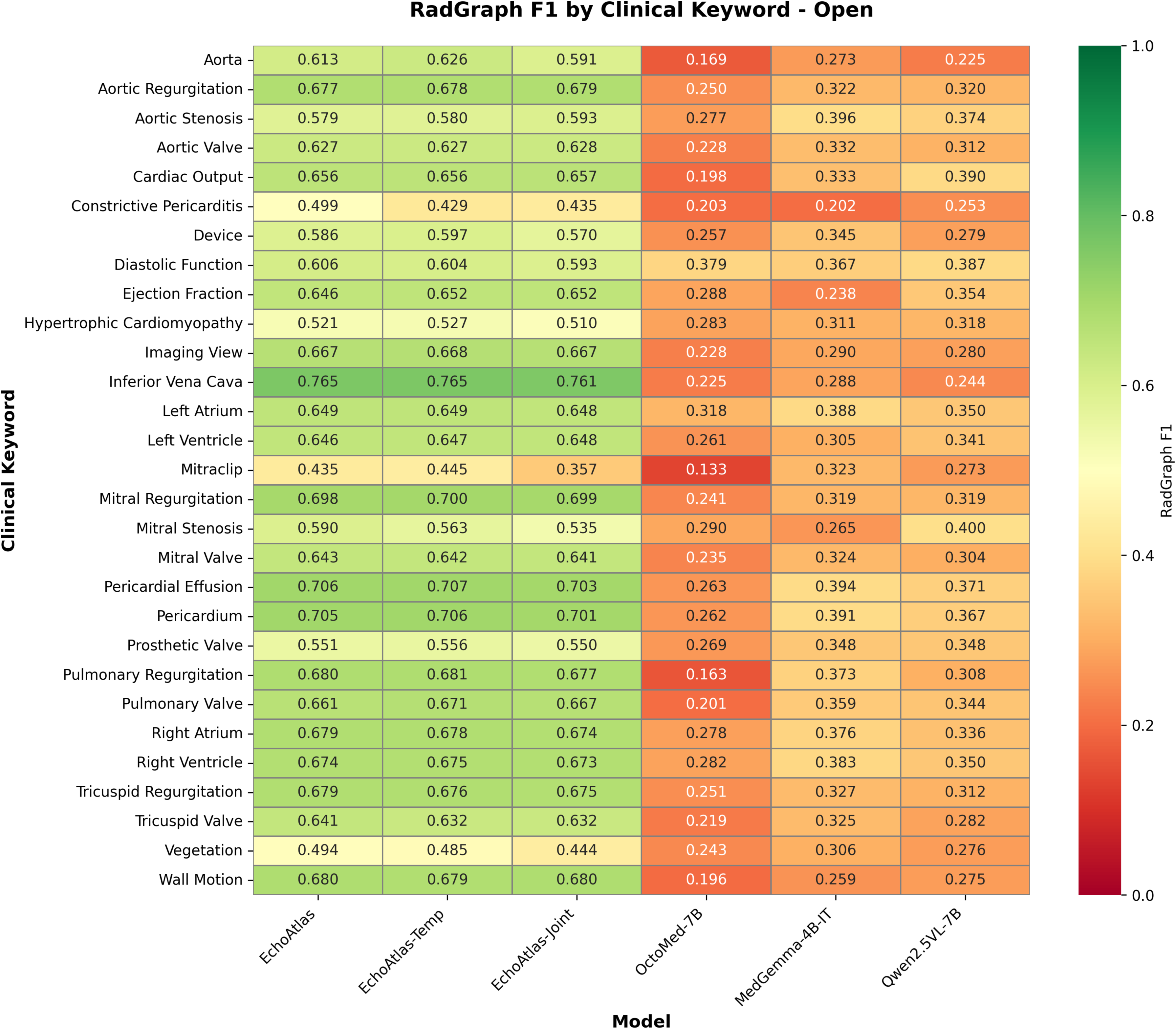
Model performance on open-ended questions stratified by clinical keyword. Heatmap displaying RadGraph F1 values (color scale 0–1; green indicates higher performance). EchoAtlas achieved robust performance (>0.5) across a majority of categories and outperformed all comparison models (OctoMed-7B, MedGemma-4B-IT, and Qwen2.5VL-7B) in every category.

**Supplementary Figure 4.**
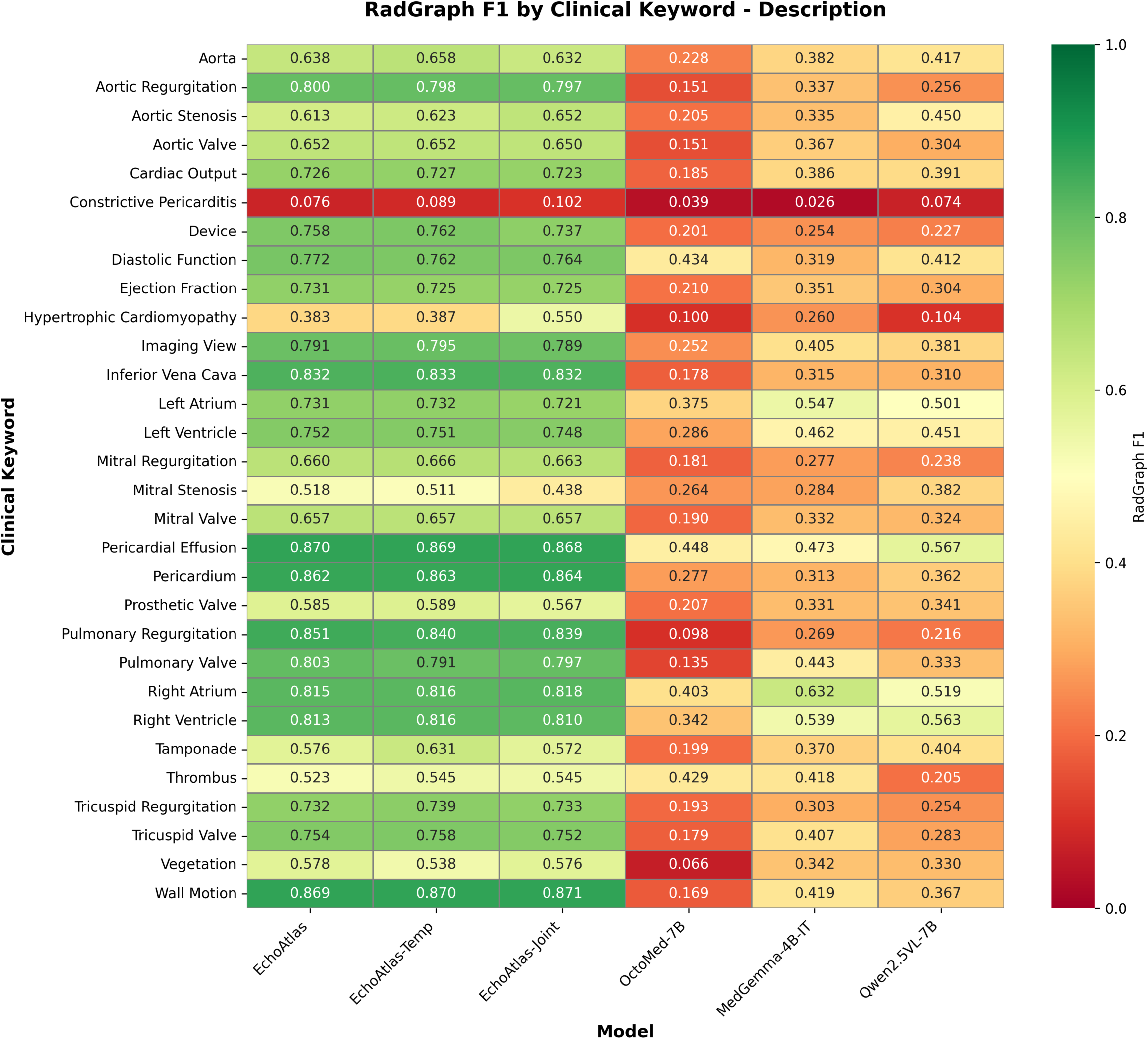
Model performance on Description questions stratified by clinical keyword. Heatmap displaying RadGraph F1 values (color scale 0–1; green indicates higher performance). EchoAtlas achieved robust performance (>0.5) across a majority of categories and outperformed all comparison models (OctoMed-7B, MedGemma-4B-IT, and Qwen2.5VL-7B) in every category.

**Supplementary Figure 5.**
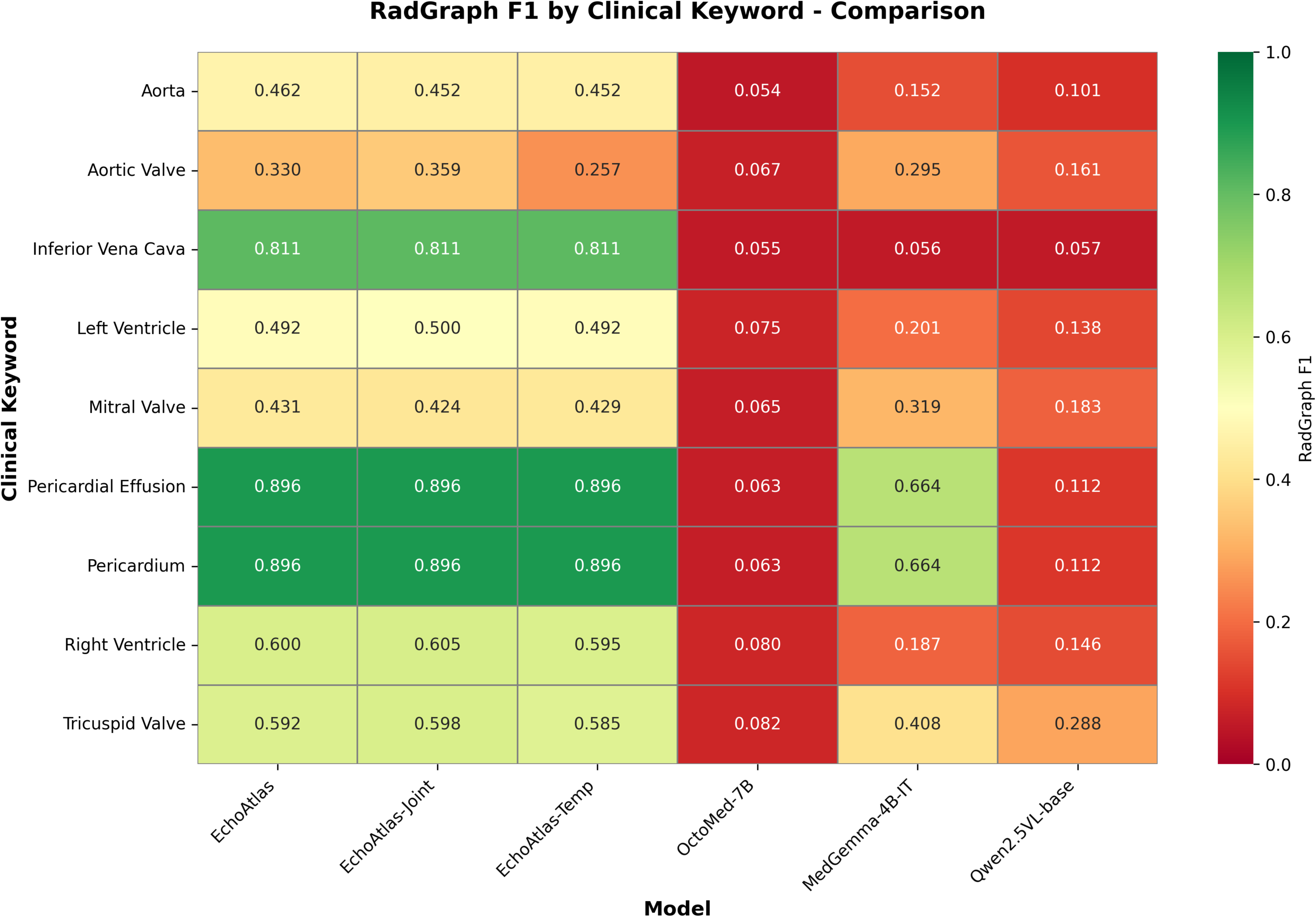
Model performance on Comparison questions stratified by clinical keyword. Heatmap displaying RadGraph F1 values (color scale 0–1; green indicates higher performance). EchoAtlas achieved robust performance (>0.5) across a majority of categories, except for comparing aorta (0.462), aortic valve (0.330), left ventricle (0.492) and mitral valve (0.431). EchoAtlas outperformed all comparison models (OctoMed-7B, MedGemma-4B-IT, and Qwen2.5VL-7B) in every category. Note aortic valve values represent results after correcting the ground truth.

**Supplementary Figure 6.**
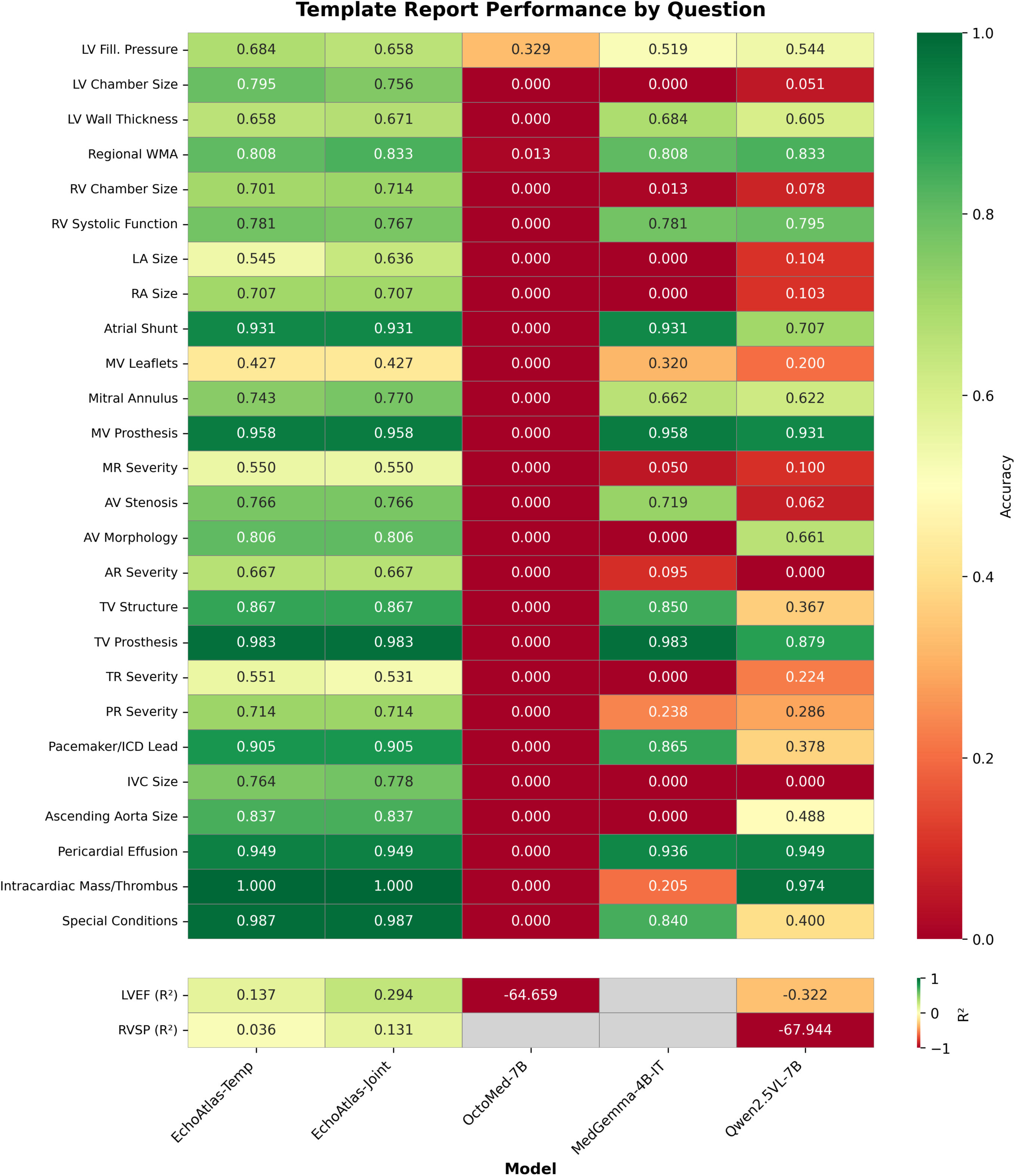
Heatmap of Template Report performance across five models. The upper panel shows per-question accuracy (color scale 0–1) for 26 structured multiple-choice templates covering LV function (LV Fill. Pressure, LV Chamber Size, LV Wall Thickness, Regional WMA), right ventricular (RV) function (RV Chamber Size, RV Systolic Function), atrial anatomy (LA Size, RA Size, Atrial Shunt), valve morphology and disease severity (MV Leaflets, Mitral Annulus, MV Prosthesis, MR Severity, AV Stenosis, AV Morphology, AR Severity, TV Structure, TV Prosthesis, TR Severity, PR Severity), cardiac devices (Pacemaker/ICD Lead), vascular structures (IVC Size, Ascending Aorta Size), and other findings (Pericardial Effusion, Intracardiac Mass/Thrombus, Special Conditions). The lower panel shows R² for two continuous measurements: left ventricular ejection fraction (LVEF R²) and right ventricular systolic pressure (RVSP R²), with gray cells indicating insufficient valid numeric predictions. Models are EchoAtlas-Temp, EchoAtlas-Joint, OctoMed-7B, MedGemma-4b-it, and Qwen2.5VL-7B. Abbreviations: WMA, wall motion abnormality; MV, mitral valve; MR, mitral regurgitation; AV, aortic valve; AR, aortic regurgitation; TV, tricuspid valve; TR, tricuspid regurgitation; PR, pulmonary regurgitation; IVC, inferior vena cava; ICD, implantable cardioverter-defibrillator; LVEF, left ventricular ejection fraction; RVSP, right ventricular systolic pressure.

**Supplementary Figure 7.**
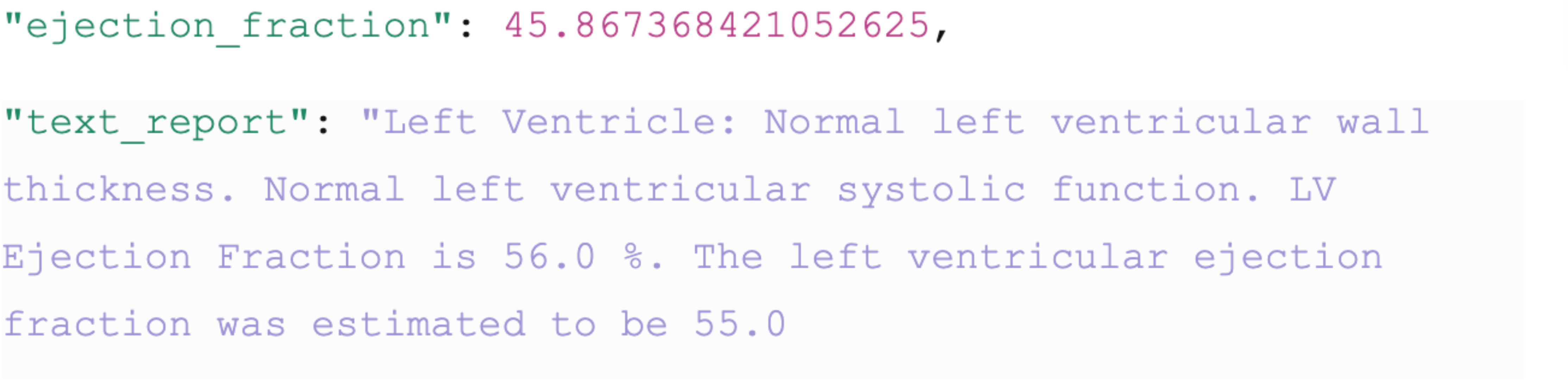
EchoPrime evaluation. During evaluations, we noticed that EchoPrime often generates contradictory values in the predicted metrics and the retrieved text, as shown in the figure. The metric predicted for ejection fraction is 45.87%, which would be categorized as mild LV systolic dysfunction, whereas the text reports it as 55-56%, as well as explicitly suggesting normal left ventricular systolic function. We found ∼0.44 accuracy when prioritizing the report and 0.48 with the metrics taking priority. The latter is reported in the main text.

**Supplementary Figure 8.**
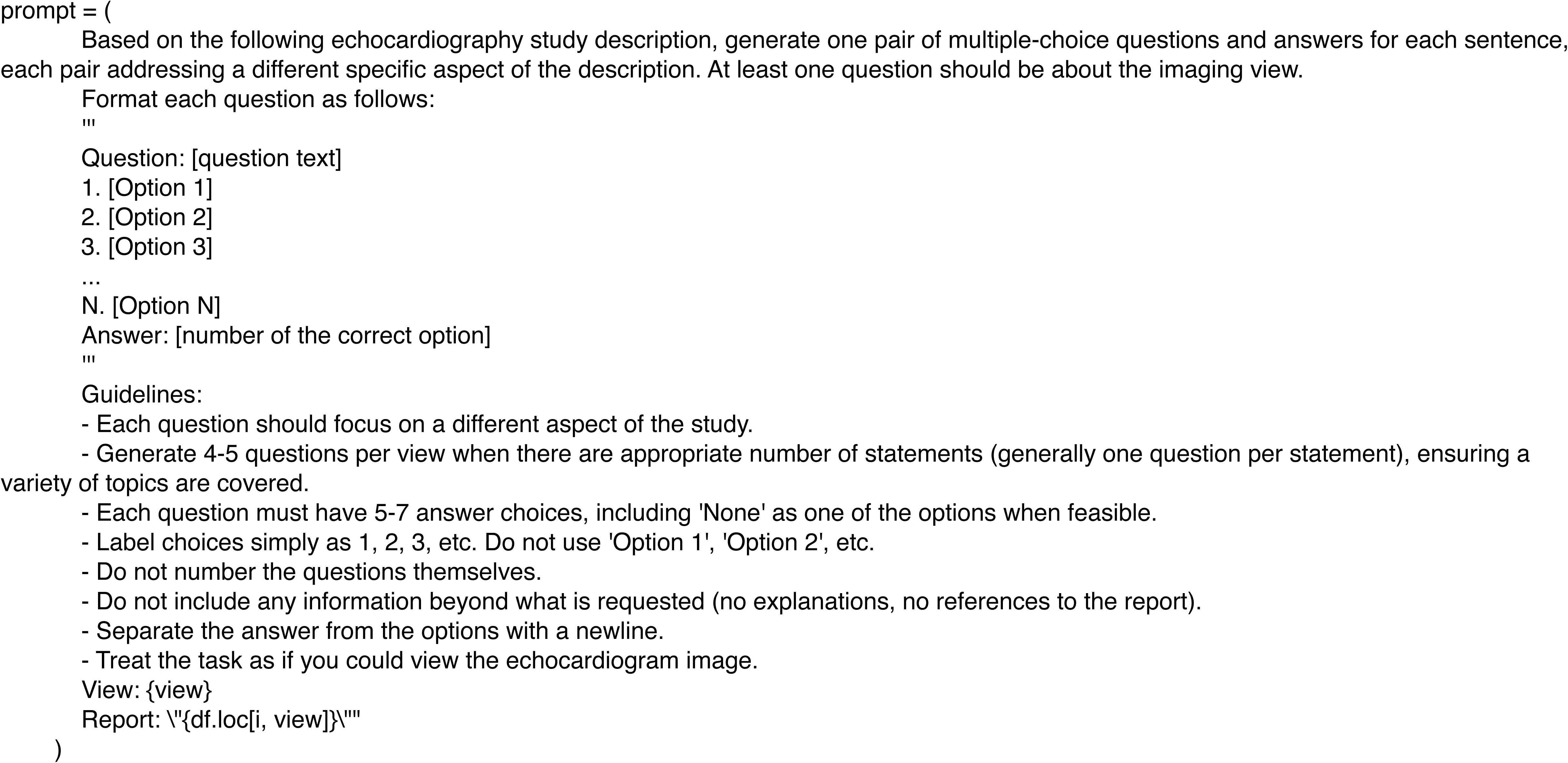
The prompt used for evaluating EchoPrime using GPT-5 (reasoning_effort=high). GPT-5 is instructed to extract the answer to the question only based on the provided (and the natural deductions from) information generated by EchoPrime. Metric values directly predicted get the priority.

**Supplementary Figure 9.**
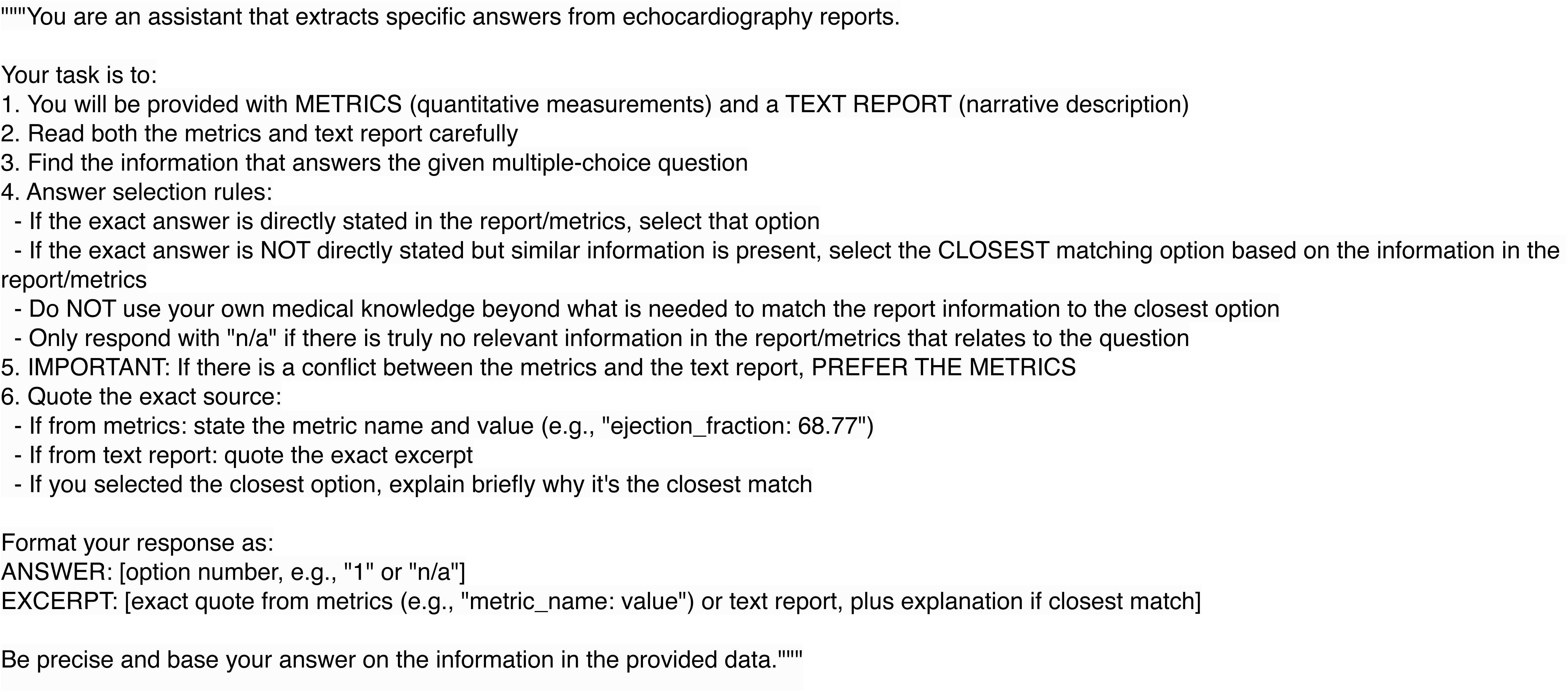

**Supplementary Table 1.** Distribution of Question Formats.

| Supplementary Table 1. Distribution of Question Formats |  |  |  |  |  |
| --- | --- | --- | --- | --- | --- |
| Format | Total Count | Train | Val | Test | Example |
| MCQ | 3,977,593 (30.9%) | 3,884,339 (30.9%) | 29,502 (28.8%) | 63,752 (30.9%) | "What is the severity of MR? 1. None, 2. Mild, 3. Moderate, 4. Severe" |
| Close | 2,242,011 (17.4%) | 2,188,688 (17.4%) | 19,177 (18.7%) | 34,146 (16.5%) | "Does hypertrophic cardiomyopathy present in this study?" |
| Open | 2,240,511 (17.4%) | 2,181,464 (17.4%) | 19,408 (18.9%) | 39,639 (19.2%) | "Is there left ventricular hypertrophy?" |
| Description | 2,126,484 (16.5%) | 2,077,126 (16.5%) | 16,906 (16.5%) | 32,452 (15.7%) | "Describe the aortic valve morphology" |
| Comparison | 2,119,887 (16.5%) | 2,070,503 (16.5%) | 15,830 (15.4%) | 33,554 (16.3%) | "What's the difference of the left ventricle between the current and prior A4C views?" |
| Measurement | 134,393 (1.0%) | 131,512 (1.0%) | 1,038 (1.0%) | 1,843 (0.9%) | "What is the Left Ventricular Cardiac Index in this study?" |
| Reasoning | 31,695 (0.2%) | 31,045 (0.2%) | 132 (0.1%) | 518 (0.3%) | "Explain why you suspect severe aortic stenosis" |
| Template Report | 7,088 (0.1%) | 6,056 (0.0%) | 474 (0.5%) | 558 (0.3%) | A combination of 26 MCQ and 2 Measurement questions, see Supplementary Table 2 Template Questions. |
| Total | 12,879,662 (100.0%) | 12,570,733 (97.6%) | 102,467 (0.8%) | 206,462 (1.6%) |  |

**Supplementary Table 2.**
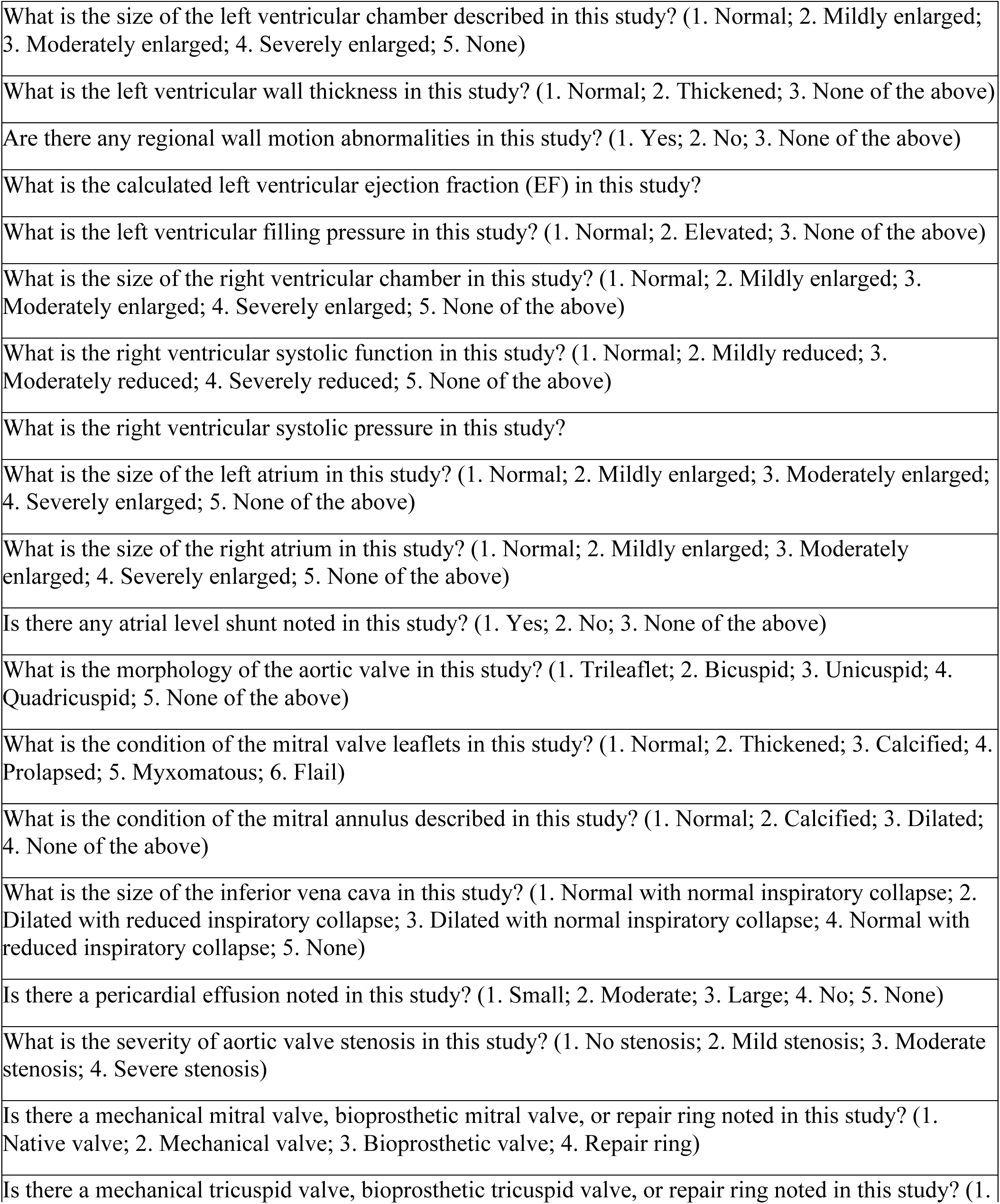
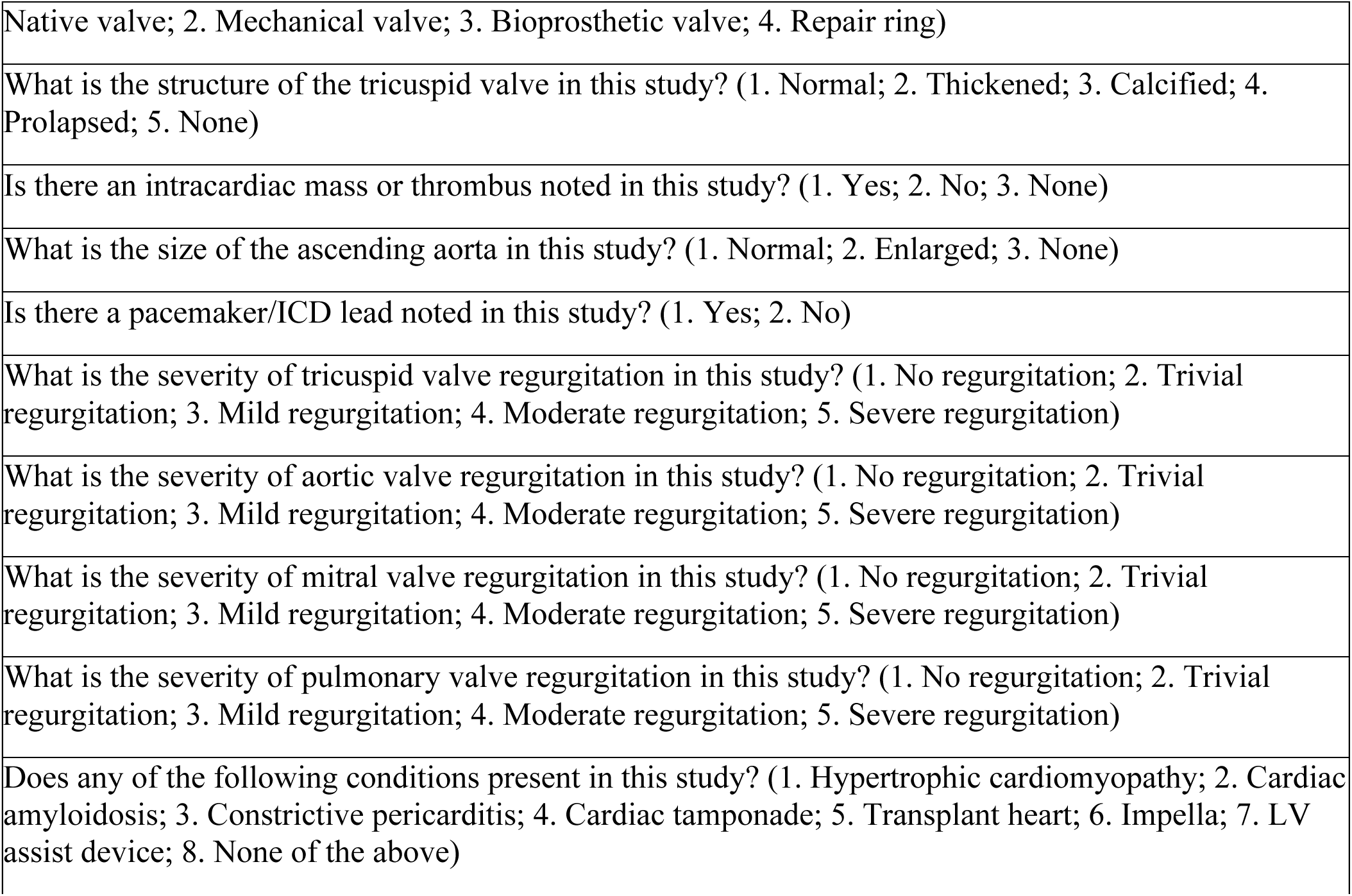
Template Questions.

**Supplementary Table 3:**
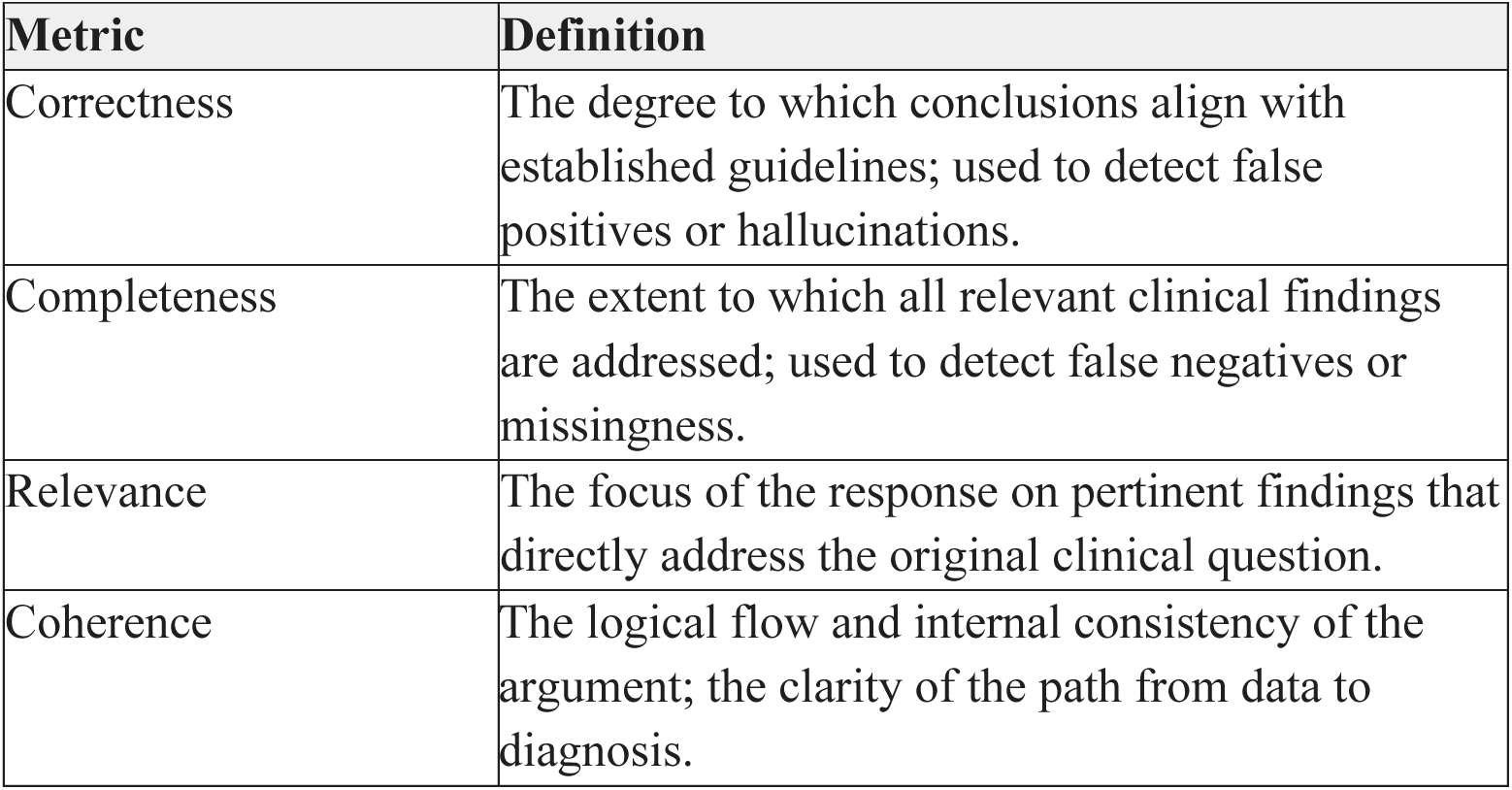
Reasoning Quality Metrics.

| Supplementary Table 3: Reasoning Quality Metrics |  |
| --- | --- |
| Metric | Definition |
| Correctness | The degree to which conclusions align with established guidelines; used to detect false positives or hallucinations. |
| Completeness | The extent to which all relevant clinical findings are addressed; used to detect false negatives or missingness. |
| Relevance | The focus of the response on pertinent findings that directly address the original clinical question. |
| Coherence | The logical flow and internal consistency of the argument; the clarity of the path from data to diagnosis. |

**Supplementary Table 4:** Error Categories for Model Performance Analysis.

|  |  |  |
| --- | --- | --- |
| Error Category | Description | Example |
| Model Error | Model clearly wrong, answer is unambiguous | Model says "Normal" when obvious severe stenosis present |
| Ambiguous Ground Truth | Reasonable disagreement is possible between experts | Mild vs. moderate severity is subjective at boundaries |
| Label Noise | Ground truth appears incorrect based on video evidence | Report says "moderate" but video shows severe pathology |
| View Mismatch | The question asks about structure not visible in view | Asking about tricuspid valve in parasternal long-axis view |
| Poor Image Quality | Video quality prevents a reliable assessment | Suboptimal acoustic windows obscure structures |

